# Disorganization of language and working memory systems in frontal versus temporal lobe epilepsy

**DOI:** 10.1101/2021.02.19.21251501

**Authors:** Lorenzo Caciagli, Casey Paquola, Xiaosong He, Christian Vollmar, Maria Centeno, Britta Wandschneider, Urs Braun, Karin Trimmel, Sjoerd B. Vos, Meneka K. Sidhu, Pamela J. Thompson, Sallie Baxendale, Gavin P. Winston, John S. Duncan, Danielle S. Bassett, Matthias J. Koepp, Boris C. Bernhardt

## Abstract

Cognitive impairment is a common comorbidity of epilepsy, and adversely impacts people with both frontal lobe epilepsy (FLE) and temporal lobe epilepsy (TLE). While the underlying neural substrates in TLE have been extensively investigated, functional imaging studies in FLE are scarce. In this study, we profiled cognitive dysfunction in FLE, and directly compared FLE and TLE patients to establish commonalities and differences. We investigated 172 adult participants (56 with FLE, 64 with TLE, and 52 controls), using neuropsychological tests and four functional MRI tasks probing the neural correlates of expressive language (verbal fluency, verb generation) and working memory (verbal and visuo-spatial). Patient groups were comparable in disease duration and anti-epileptic drug load. We devised a multiscale approach to map the landscape of brain activation and deactivation during cognition, and track reorganization in FLE and TLE. Voxel-based analyses were complemented with profiling of task effects (i) across intrinsic functional systems, and (ii) along the principal functional connectivity gradient, which encodes a continuous transition from lower-level sensory to higher-order transmodal brain areas. We show that cognitive impairment in FLE is associated with reduced activation across attentional and executive systems, and reduced deactivation of the default mode system, indicative of a large-scale disorganization of task-related recruitment. Functional abnormalities in FLE were modulated by disease load. Patterns of dysfunction in FLE were broadly similar to those in TLE, but some traits were syndrome-specific: altered default-mode deactivation was more prominent in FLE, while impaired recruitment of posterior language areas during a task with semantic demands was more marked in TLE. Our study elucidates neural processes underlying language and working memory impairment in FLE, identifies shared and syndrome-specific alterations in the two most common focal epilepsies, and sheds light on system behavior that may be amenable to future remediation strategies.

## INTRODUCTION

Frontal lobe epilepsy (FLE), the second most common focal epilepsy syndrome after temporal lobe epilepsy (TLE), is frequently drug-resistant and MRI-negative.^1–3^ Cognitive dysfunction is common in both FLE and TLE, and adversely impacts quality of life and psychosocial functioning.^4^ Impaired episodic memory and semantic knowledge are common in TLE, though dysexecutive traits frequently coexist.^5–7^ In contrast, FLE has a less established cognitive signature. Multiple domains may be affected, including dexterity, attention, working memory, verbal fluency, executive functions, and episodic memory.^8–12^ Whether cognitive profiles in FLE and TLE may be distinct remains controversial, and several investigations concluded that these syndromes cannot be discriminated on cognitive measures.^9, 13, 14^ It is however suggested that episodic memory impairment is more profound in TLE, while executive functions may be more affected in FLE.^8, 15–17^

Task-based functional MRI (fMRI) probes the neural correlates of cognitive dysfunction in epilepsy. In TLE, altered activation and connectivity of parietal and mesiotemporal areas underlie working memory impairment,^18, 19^ while language fMRI studies indicate altered fronto-temporal activation and connectivity, with complex intra- and inter-hemispheric reorganization.^20–26^ In contrast, only few studies^27^ investigated FLE. Pediatric FLE patients had reduced fronto-temporo-parietal connectivity on a working memory fMRI task, but no substantial alteration in regional activation.^28^ Previously, we reported enhanced fronto-temporal activation during episodic memory encoding in adult FLE patients, along with reduced mesiotemporal activation in those with poorer memory.^29^ Abnormal activity of motor and posterior parietal cortices during a motor coordination task may underlie impaired dexterity.^30^ Overall, a comprehensive overview of the neural substrates of cognitive dysfunction in FLE is lacking.

Here, we aimed to characterize the functional neuroanatomy of expressive language and working memory, cognitive functions reliant on frontal lobe processing,^31, 32^ in individuals with drug-resistant FLE who underwent neuropsychological tests and four fMRI tasks. We compared patients with FLE to (i) healthy controls, and to (ii) a “patient control group” of individuals with TLE, comparable for epilepsy duration and anti-seizure medication (ASM) load, which allowed establishing shared and syndrome-specific traits.

We devised a multiscale functional mapping scheme to investigate the landscape of competing brain activation and deactivation during cognition,^33–35^ and capture disease-related reorganization. We complemented traditional voxel-based activation analyses by profiling task effects (i) across established cognitive systems,^36^ and (ii) in relation to the principal gradient of intrinsic functional connectivity.^37, 38^ The gradient describes a continuous transition of neural function, with unimodal sensory areas at one end, and high-order transmodal regions at the opposite end. As such, it recapitulates established models of cortical hierarchy,^39^ and offers a formal framework to characterize organizational aspects of cognitive activity, as exemplified by recent work in healthy adults^40–42^ and people with TLE engaged in a pattern separation task.^43^ By conveying regional, systems-level and global viewpoints on the neural signatures of cognitive dysfunction in epilepsy, our innovative approach proves sensitive to both localized and higher-order abnormalities.

We anticipated expressive language and working memory impairment in FLE. We hypothesized that such impairment would be underpinned by (i) reduced activation of areas engaged during task execution, i.e., “task-positive” regions, (ii) reduced deactivation of default-mode areas (DMN), i.e., “task-negative” regions, and (iii) global disorganization of cognitive network recruitment, as quantified via gradient analysis. We also hypothesized that, based on the proximity to the epileptic focus, (i) frontal and systems-level working memory abnormalities may be more prominent in FLE than TLE, (ii) language-related activation of frontal areas would be worse in FLE, and (iii) engagement of temporal language areas would be worse in TLE. We also aimed to corroborate the neurobehavioral validity of our fMRI tasks by correlating imaging patterns with neuropsychological and task performance measures. Finally, we explored associations between cognitive network alterations and clinical characteristics considered as measures of disease load, such as age at onset and epilepsy duration, and replicated our main FLE findings in a more homogeneous patient subgroup with frontal cortical dysplasia.

## MATERIALS AND METHODS

### Participants

This study investigated 172 participants recruited from 2007 to 2013: 120 drug-resistant patients under surgical consideration, of whom 56 had drug-resistant FLE (29 female; 30/26 left-/right-sided FLE) and 64 had drug-resistant TLE (44 female; 34/30 left-/right-sided TLE), and 52 healthy controls (32 female) with no family history of epilepsy and no neurological or psychiatric diagnosis. Demographic and clinical characteristics are provided in Table 1; further details are available in the Supplementary Methods. Written informed consent was obtained from all participants according to the standards of the Declaration of Helsinki. Participant recruitment was approved by the University College London Institute of Neurology and University College London Hospitals Research Ethics Committee. Exclusion criteria were non-proficiency in written and spoken English, MRI contraindications, pregnancy, and inability to give informed consent.

**Table 1.** Demographic, clinical and neuropsychological data.

|  | **CTR**<br>**(n=52)** | **FLE**<br>**(n=56)** | **TLE**<br>**(n=64)** | **Test**<br>**statistic**<br>**(*F*, *H* or $\chi^2$)** | ***P* value** | ***Post-hoc* tests**<br>**(Bonferroni**<br>**corrected)** |
| --- | --- | --- | --- | --- | --- | --- |
| **Age at scan**<br>[years, mean (SD)] | 34.1<br>(10.4) | 33.4<br>(10.2) | 39.2<br>(10.7) | 5.6 | **0.005** | FLE/CTR: 1.00<br>FLE/TLE: **0.008**<br>TLE/CTR: 0.029 |
| **Sex (F/M)** | 30/22 | 29/27 | 44/20 | 3.7 | 0.15 |  |
| **Handedness (L/R/A)** | 6/46/0 | 5/49/2 | 10/52/2 | 3.0 | 0.59 |  |
| **Side of seizure focus (L/R)** | — | 30/26 | 34/30 | 0.0 | 1.00 |  |
| **Etiological categories**<br>(non-lesional/FCD/DNET/<br>glial tumor/other†/HS) | — | 29/13/6/<br>3/5/0 | 0/0°/0°/<br>0/0/64 | — | — |  |
| **Age of epilepsy onset**<br>[median (IQR)] | — | 10.0<br>(6.0) | 13.0<br>(11.8) | 2.3 | 0.13 |  |
| **Duration of epilepsy**<br>[median (IQR)] | — | 21.0<br>(15.0) | 22.5<br>(24.3) | 0.6 | 0.44 |  |
| **AEDs**<br>[median (IQR)] | — | 3.0<br>(1.0) | 2.5<br>(1.0) | 1.3 | 0.25 |  |
| **Topiramate/Zonisamide**<br>[yes/no] | — | 15/41 | 12/52 | 1.1 | 0.38 |  |
| **Levetiracetam**<br>[yes/no] | — | 29/27 | 41/23 | 1.9 | 0.20 |  |
| **Frontal language LI**<br>[Verbal fluency fMRI,<br>median (IQR)] | 0.79<br>(0.32) | 0.67<br>(0.56) | 0.76<br>(0.29) | 10.5 | **0.005** | FLE/CTR: **0.004**<br>FLE/TLE: 0.094<br>TLE/CTR: 0.744 |
| **Frontal language LI**<br>[Verb generation fMRI,<br>median (IQR)] | 0.81<br>(0.23) | 0.65<br>(1.15) | 0.70<br>(0.41) | 9.1 | **0.01** | FLE/CTR: **0.019**<br>FLE/TLE: 1.00<br>TLE/CTR: **0.030** |
| **IQ (NART)**<br>[mean (SD)] | 112.1<br>(8.8) | 99.7<br>(11.0) | 97.3<br>(12.3) | 24.2 | **3.3e-**<br>**09\*** | FLE/CTR: **1.0e-06**<br>FLE/TLE: 0.58<br>TLE/CTR: **1.5e-09** |
| **Letter Fluency**<br>[mean (SD)] | 18.8<br>(6.1) | 10.9<br>(4.9) | 13.2<br>(6.0) | 23.8 | **3.5e-**<br>**09\*** | FLE/CTR: **1.3e-9**<br>FLE/TLE: 0.25<br>TLE/CTR: **3.0e-06** |
| **Category Fluency**<br>[mean (SD)] | 24.0<br>(4.9) | 18.2<br>(5.9) | 17.9<br>(5.1) | 19.4 | **8.8e-**<br>**08\*** | FLE/CTR: **1.0e-06**<br>FLE/TLE: 1.00<br>TLE/CTR: **2.3e-07** |
| **Graded Naming Test**<br>[mean (SD)] | 22.0<br>(3.6) | 16.7<br>(4.3) | 14.3<br>(5.7) | 42.7 | **3.8e-**<br>**14\*** | FLE/CTR: **2.0e-06**<br>FLE/TLE: **4.4e-04**<br>TLE/CTR: **7.8e-16** |
| **Vocabulary**<br>[mean (SD)] | — | 9.1<br>(3.7) | 8.0<br>(2.8) | -1.8§/-<br>5.7§/3.0 | — | FLE/norm: 0.23<br>FLE/TLE: 0.26<br>TLE/norm: 1.2e-06 |
| **Similarities**<br>[mean (SD)] | — | 8.3<br>(3.2) | 8.5<br>(2.8) | -3.9§/-<br>4.3§/0.3 | — | FLE/norm: 9.5e-04<br>FLE/TLE: 1.00<br>TLE/norm: 1.7e-04 |
| **Digit Span**<br>[mean (SD)] | 11.4<br>(2.6) | 8.0<br>(2.9) | 8.0<br>(2.2) | 25.1 | **2.0e-**<br>**09\*** | FLE/CTR: **8.2e-09**<br>FLE/TLE: 1.00<br>TLE/CTR: **8.2e-09** |
| **Trail Making A^**<br>[mean (SD)] | 28.1<br>(8.8) | 41.4<br>(17.8) | 35.9<br>(14.7) | 10.7 | **7.3e-**<br>**05\*** | FLE/CTR: **2.7e-05**<br>FLE/TLE: 0.13<br>TLE/CTR: **0.022** |
| **Trail Making B-A^**<br>[mean (SD)] | 25.8<br>(11.2) | 69.0<br>(55.5) | 47.4<br>(36.5) | 18.6 | **1.7e-**<br>**07\*** | FLE/CTR: **3.2e-08**<br>FLE/TLE: **0.015**<br>TLE/CTR: **0.002** |
| **List Learning (A1-5)**<br>[mean (SD)] | 58.1<br>(8.2) | 49.9<br>(9.6) | 43.9<br>(10.8) | 26.0 | **1.7e-**<br>**09\*** | FLE/CTR: **1.0e-04**<br>FLE/TLE: **0.010**<br>TLE/CTR: **8.04e-11** |
| **List Learning (A6)**<br>[mean (SD)] | 12.1 (3.0) | 10.4<br>(3.4) | 8.4<br>(3.5) | 15.5 | **1.5e-**<br>**06\*** | FLE/CTR: 0.10<br>FLE/TLE: **0.002**<br>TLE/CTR: **6.4e-07** |
| **Design Learning (A1-5)**<br>[mean (SD)] | 38.8<br>(5.1) | 32.3<br>(9.6) | 31.4<br>(8.0) | 11.7 | **3.0e-**<br>**05\*** | FLE/CTR: **1.2e-04**<br>FLE/TLE: 1.00<br>TLE/CTR: **1.2e-04** |
| **Design Learning (A6)**<br>[mean (SD)] | 8.2<br>(1.4) | 6.8<br>(2.4) | 6.4<br>(2.6) | 6.8 | **0.002\*** | FLE/CTR: **0.005**<br>FLE/TLE: 1.00<br>TLE/CTR: **0.004** |
| **Verbal WM task–Verbal**<br>**Monitoring** | 100.0<br>(0.0) | 100.0<br>(7.1) | 100.0<br>(7.1) | 1.3 | **0.53\*** |  |

| [median (IQR)] |  |  |  |  |  |  |
| --- | --- | --- | --- | --- | --- | --- |
| **Verbal WM task – 2 Back**<br>[median (IQR)] | 100.0<br>(9.6) | 92.3<br>(19.2) | 84.6<br>(28.9) | 16.4 | **3.6e-04\*** | FLE/CTR: **0.014**<br>FLE/TLE: 1.00<br>TLE/CTR: **2.4e-04** |
| **Visual WM task – 0 Back**<br>[median (IQR)] | 94.7<br>(7.3) | 94.7<br>(8.7) | 90.7<br>(15.0) | 12.1 | **0.002\*** | FLE/CTR: 1.00<br>FLE/TLE: **0.027**<br>TLE/CTR: **0.004** |
| **Visual WM task – 1 Back**<br>[median (IQR)] | 90.0<br>(20.0) | 78.6<br>(38.6) | 72.9<br>(42.1) | 14.8 | **7.5e-04\*** | FLE/CTR: **0.013**<br>FLE/TLE: 1.00<br>TLE/CTR: **0.001** |
| **Visual WM task – 2 Back**<br>[median (IQR)] | 73.8<br>(37.7) | 43.1<br>(28.5) | 40.0<br>(44.6) | 22.7 | **2.1e-05\*** | FLE/CTR: **1.8e-04**<br>FLE/TLE: 1.00<br>TLE/CTR: **5.7e-05** |

Abbreviations: CTR= healthy controls; DNET= dysembryoplastic neuroepithelial tumor; FCD= suspected focal cortical dysplasia; FLE= frontal lobe epilepsy; HS= hippocampal sclerosis; IQ= intelligence quotient; IQR= interquartile range; LI= laterality index; NART= National Adult Reading Test; norm= normative ranges (Wechsler scale); SD = standard deviation; TLE= temporal lobe epilepsy; WM= working memory. Categorical variables were compared with Fisher's exact test ( $\chi^2$ statistic is reported). Continuous normally/non-normally distributed demographic variables are reported as mean (SD)/median (IQR), and were compared via ANOVA ( $F$ statistic)/Kruskal-Wallis test ( $H$ statistic), respectively. Pairwise deletion was applied in case of missing data. Age at onset (and, consequently, duration) could not be accurately estimated in 5 patients with FLE. For details about computation of language laterality indices, see *Supplementary Methods*.
Neuropsychological test scores were compared via ANCOVA, using age and sex as covariates, except for: (i) digit span scores, already adjusted for age (as per Wechsler score scales), for which ANCOVA with sex as covariate was used; (ii) Vocabulary and Similarities, for which: one-sample $t$ -tests (§) assessed deviation of FLE and TLE data from normative scores as provided by the Wechsler Adult Intelligence Scale (*Supplementary Methods*), and ANCOVA with sex as covariate was used to compare FLE to TLE.
† One patient with possible periventricular nodular heterotopia, and five with miscellaneous MRI abnormalities (described in detail in the *Supplementary Methods*); °Hippocampal sclerosis coexisted with DNET in three patients (one left/two right), and with a possible FCD in one patient (right). ^ Statistics for Trail Making Test A and B-A were carried out on log-transformed data, but raw data are provided in the table to ensure comparability with published literature. Working memory task performance measures were compared via Kruskal-Wallis tests. \* $P$ -values for group comparisons across neuropsychological and task performance measures were adjusted for multiple comparisons via the False Discovery Rate (FDR) procedure. *Post-hoc* tests are all Bonferroni-corrected.

Groups were comparable for handedness and (binary) sex, but not for age, which was used as a covariate in all group analyses. Patient groups did not differ in age at onset and duration of epilepsy, number of ASMs, and usage of levetiracetam or topiramate/zonisamide, which more favorably or unfavorably influence cognitive network activity compared to other common ASMs, respectively.^44, 45^ FLE was heterogeneous in terms of etiology and MRI findings (Table 1, Supplementary Material). We thus conducted sensitivity analyses on a subgroup with a more homogeneous etiology (focal cortical dysplasia; FLE-FCD, n=13). Moreover, we separately studied left and right FLE subgroups.

### Neuropsychological data

Participants underwent standardized neuropsychological tests^46^ providing measures of general intellectual level (IQ), working memory, letter and category fluency, naming, psychomotor speed and executive function (mental flexibility), verbal and visuo-spatial learning and recall. Verbal reasoning and comprehension scores were available for patient groups. Test details are provided in the Supplementary Methods.

### Imaging data acquisition and fMRI tasks

All imaging data were acquired on the same GE Signa HDx 3T MRI scanner at the Epilepsy Society MRI Unit, Chalfont St Peter, Buckinghamshire. Data acquisition is described in the Supplementary Methods. One visuo-spatial and one verbal fMRI paradigm assessed working memory. During the visuo-spatial (Dot Back) task, dots appeared in four possible locations on a screen. Participants were instructed to move a joystick to the position of the currently presented dot (0Back), or to the position of the dot displayed one (1Back) or two presentations earlier (2Back).^47^ There were five 30s blocks for each condition in pseudo-random order, intermixed with 15s of cross-hair fixation. During the verbal working memory task, single concrete nouns were displayed every 3s within 30s blocks. Participants responded upon display of a given control word (active control condition), or upon recurrence of a word displayed two presentations earlier (2Back working memory). There were five 30s blocks per condition, intermixed with 15s of cross-hair fixation. Two tasks probed expressive language. During verbal fluency fMRI, participants covertly generated words beginning with a visually-presented letter (A/D/E/S/W; one letter per block, five 30s blocks), alternating with 30s blocks of cross-hair fixation.^48^ During the verb generation task, subjects covertly generated verbs associated with a visually-displayed noun (“Generate”), or subvocally repeated a visually-displayed noun (“Repeat”). There were four 30s blocks per condition and four cross-hair fixation blocks.^49^

### Statistical analysis of clinical and neuropsychological data

Data were analyzed using *R*-3.6.1 and SPSS-27. For demographics, we used Fisher’s exact test, one-way ANOVA and Kruskal-Wallis tests for categorical, continuous parametric and nonparametric variables, respectively. Neuropsychological data were compared via ANCOVA, covarying for age and sex. Comparisons against published norms were attained with one sample *t-*tests (details in the Supplementary Methods). Working memory task performance measures were not normally distributed, and were compared via Kruskal-Wallis tests. Across cognitive domains, we corrected for multiple comparisons via the false discovery rate (FDR) procedure.^50^ *Post-hoc* tests were Bonferroni-corrected.

### Functional MRI data: pre-processing and voxel-based statistics

Functional imaging data were analyzed with SPM12 (http://www.fil.ion.ucl.ac.uk/spm/). Preprocessing included realignment, normalization to a scanner- and acquisition-specific echo-planar imaging template in Montreal Neurological Institute (MNI) space, resampling to 3×3×3mm isotropic voxels, and smoothing with a Gaussian kernel of 8×8×8mm full-width at half-maximum.^51^ Individual-level condition-specific effects were derived via general linear models. Task conditions were modelled as 30s blocks and convolved with the canonical hemodynamic response function. For verbal fluency fMRI, we created activation contrasts associated with generating words. For verb generation fMRI, we subtracted word repetition from word generation. For verbal working memory fMRI, we subtracted verbal monitoring from the 2Back working memory condition. For visuo-spatial working memory fMRI, we contrasted the condition with low working memory demand against the active control condition (1–0Back), and directly compared activation for high and low working memory demand (2– 1Back). Voxel-wise contrast estimates (β weights) were computed with six motion parameters as confound regressors. Scans with a mean framewise displacement >0.5mm were discarded from further analysis.^52^ Additional quality checks and final participant numbers are provided in the Supplementary Methods.

Group analyses were conducted with nonparametric permutation tests, using SnPM13^53^ (http://www.nisox.org/Software/SnPM13/). One-sample permutation *t-*tests assessed effects of each task condition per group. Following exploratory permutation-based *F-*tests, group differences were assessed via two-sample permutation *t*-tests, all with 10000 permutations. Statistical significance was set at two-tailed p<0.05, voxel-wise corrected for familywise-error rate (FWE)^54^ within pre-specified language, working memory and DMN (“task-negative”) regions of interests (ROIs; Supplementary Fig. 1). All comparisons included age and sex as covariates; comparisons of FLE versus TLE included side of the epileptic focus as additional covariate. Further details regarding group comparisons are outlined in Table 2. ROI definition is discussed in the Supplementary Methods.

**Table 2.**
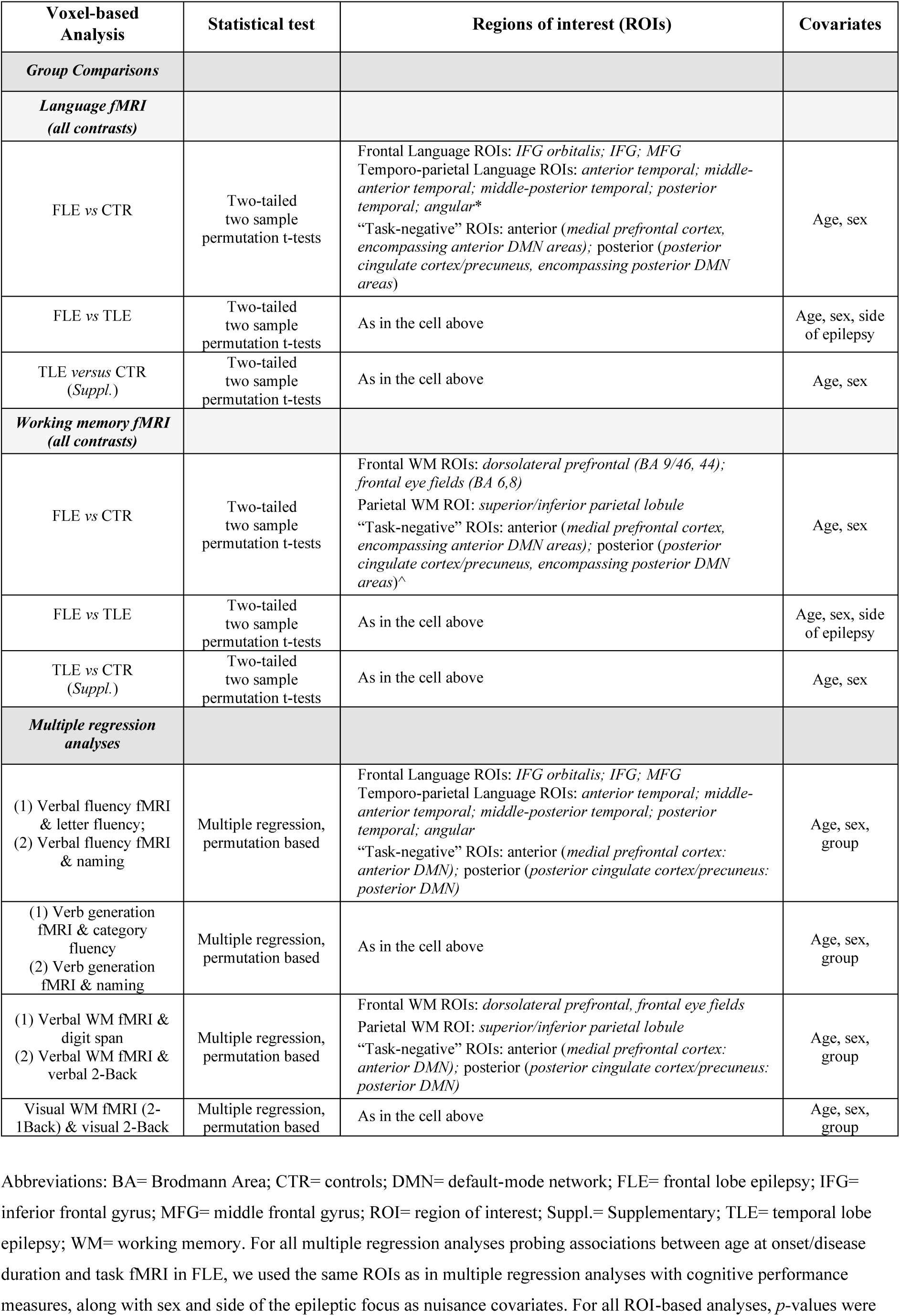
Voxel-based group comparisons and association analyses: further details.

### Functional MRI data: effects on canonical systems and principal gradient

Voxel-based analyses elucidate task effects and group differences at a regional level. Pursuant to a higher-order, ensemble view on cognition-related brain activity, we quantified task-effects across seven intrinsic functional systems^36^ (Figure 1): visual, somatomotor, dorsal attention, salience (or ventral attention), (para)limbic, cognitive control (frontoparietal), and DMN. For each task contrast, we extracted β weights from all parcels of the *Schaefer* brain atlas^55^ (200 ROI scale, MNI space) via FSL-6.0.2, averaged β weights across ROIs belonging to a given system, and adjusted the latter for age and sex. Profiling of task effects along the principal functional gradient was conducted in surface space^37, 56^ (Supplementary Methods). The gradient was computed from resting-state fMRI data of 100 Human Connectome Project (HCP) participants via non-linear dimensionality reduction of surface-registered functional connectivity metrics.^57, 58^ HCP acquisition and preprocessing have been detailed elsewhere.^59^

**Figure 1.**
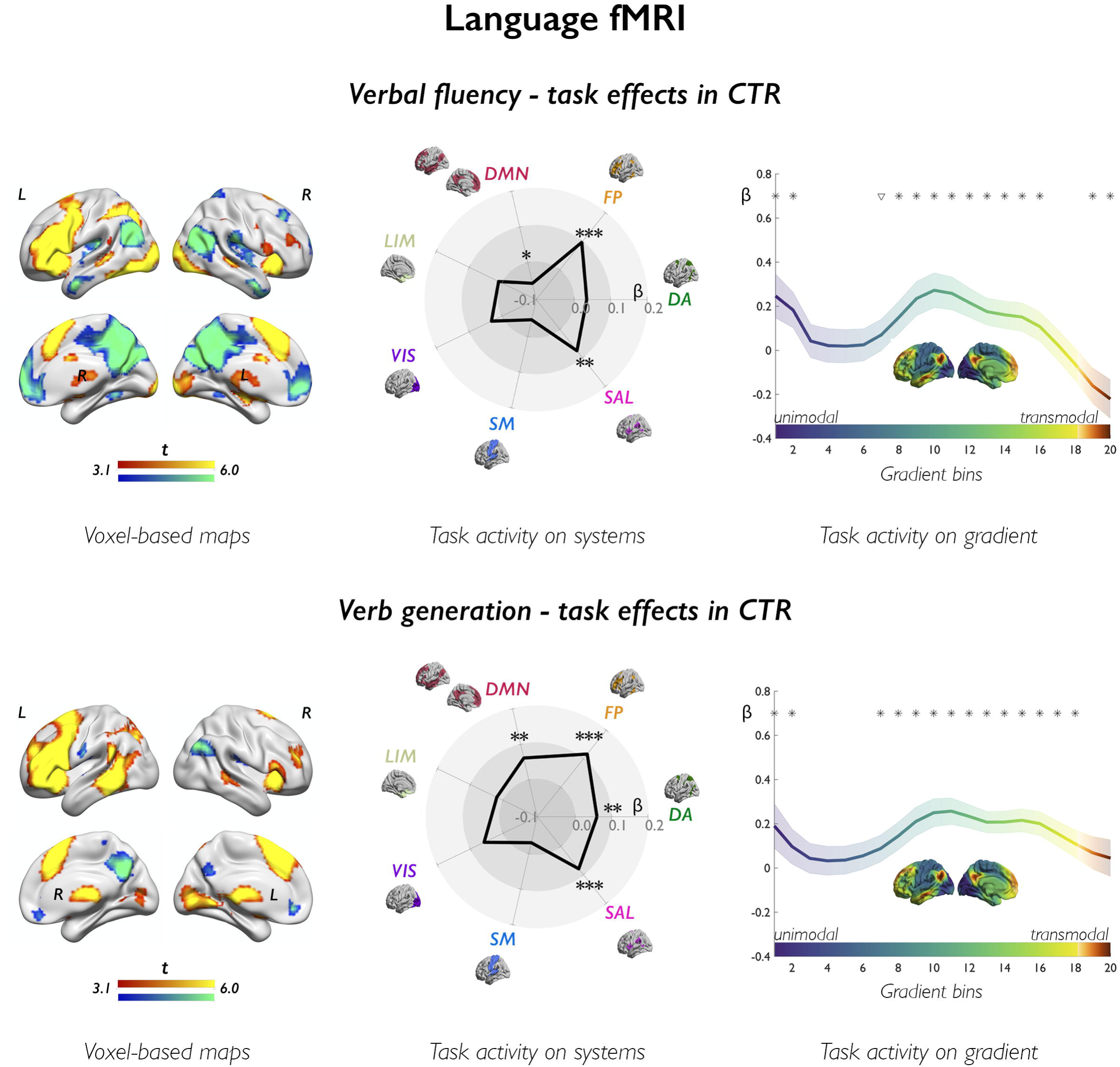
Language fMRI: task effects in controls. The upper and lower panels show task effects in controls (CTR) for verbal fluency and verb generation fMRI, respectively. Left-sided panels show voxel-based activation (warm colors) and deactivation (cold colors) in controls, as derived from one-sample *t*-tests. Brain renders show maps at p<0.001, with an extent threshold of 10 voxels applied for display purposes; color bars indicate t-score scales. The spider plots (middle panels) show mean task-related effects, parameterized as β weights (contrast estimates) across 7 canonical systems, where DA: dorsal attention; FP: cognitive (frontoparietal) control; DMN: default-mode network; LIM: (para)limbic; VIS: visual; SM: somatomotor; SAL: salience (ventral attention); ***, *p*_FDR_<0.01; **, *p*_FDR_<0.05; *, uncorrected *p*<0.05. The upper and lower right-sided panels show task-related effects (β weights) stratified along the principal gradient, which depicts a continuous coordinate system running from unimodal (dark blue) areas at one end, to high-order transmodal (sienna) regions at the other end; its left lateral and midline views are provided in the middle of each plot, using the same color scale as in curves of gradient-stratified task effects. The gradient was discretized into 20 consecutive, equally-sized bins (*x* axis), with task-related signal (β weights; *y* axis) computed per bin; shaded areas refer to 95% confidence intervals of mean effects at each bin; *, *p*_FDR_<0.05; ∇, uncorrected *p*<0.05.

The gradient was discretized into 20 equally-sized bins, as previousy;^40, 56^ cortical locations were assigned to each bin, with mainly sensory or motor regions assigned to the 1^st^ bin, and transmodal regions assigned to the 20^th^ bin. For each participant and task contrast, we derived average β weights in each bin via a sliding window approach,^60^ and adjusted them for age and sex.

### Functional MRI data: statistics for system and gradient analyses

In controls, one-sample permutation *t*-tests assessed task effects per system or gradient bin. We computed deviation (Z) scores to determine the atypicality of effects in patients [Z*_pat_* = (Act*_pat_* −μ*_CTR_*)/σ*_CTR_*], where μ*_CTR_* and σ*_CTR_* correspond to the mean and standard deviation of systems-level or bin-wise β weight in controls for a given task contrast.^61, 62^ For each system or gradient bin, Z-score deviations from zero in patients were assessed with two-tailed, permutation-based one-sample *t-*tests. FLE and TLE were compared via permutation-based two-tailed two-sample *t-*tests. For gradient analyses, group comparisons also assessed global differences between curves of gradient-stratified task effects (areas between curves; AbC), using a nonparametric permutation test based on functional data analysis techniques^63^ (FDA; Supplementary Methods). We used 10000 permutations for all tests, and report Cohen’s *d* effect sizes. *P*-values were FDR-adjusted for number of systems or gradient bins; comparisons reaching uncorrected *p<*0.05 (*p*_unc_) and related effect sizes are reported for completeness. Group comparisons constrained to the left-hemispheric sections of the seven systems are also reported for language tasks, given the predominant left lateralization of language processing. Follow-up analyses of effects across DMN and cognitive control system subdivisions, derived from a more fine-grained 17-system parcellation,^36^ are shown in the Supplementary Results.

### Correlation of functional MRI data with cognitive performance and clinical variables

Across scales, we assessed correlations of task effects with cognitive performance in all participants^22, 43^ using permutation-based analyses, each entailing 10000 permutations. Voxel-based regressions were conducted with SnPM13; age, sex and group were nuisance covariates. Associations between cognitive scores and fMRI metrics were explored within language, working memory and task-negative ROIs (Table 2). Effects are reported at two-tailed, voxel-wise *p*_FWE_<0.05, unless otherwise stated. For correlations between task effects across systems or on the gradient, parameterized as age- and sex-adjusted β weights, and cognitive performance measures, we employed permutation-based two-tailed product-moment correlations. For verbal and visual working memory task performance measures, which were skewed, we employed permuted rank correlations with 10000 permutations. Correlations between fMRI activation and clinical variables, such as age at onset and disease duration,^22^ were separately computed in FLE and TLE, to disentangle syndrome-specific effects. In SnPM13, voxel-based regressions used sex and side of seizure focus as covariates. Statistical significance was established using the same ROIs as above. For correlations between clinical variables and task effects across systems or gradient, we used two-tailed, permutation-based product-moment correlations.

### Data availability

Data supporting the findings of this study are available from the corresponding authors upon reasonable request.

Analytic code will be available at https://github.com/lcaciagl/Language_WM_FLE_vs_TLE upon manuscript acceptance for publication.

## RESULTS

### Neuropsychological data and fMRI task performance

Patients with FLE differed from controls and/or published norms for most cognitive measures (all *p*_FDR_<0.001; Table 1). Compared to patients with TLE, those with FLE scored higher on naming, verbal learning, and verbal recall tests, and lower on a mental flexibility test (*post-hoc* p<0.05, Bonferroni-corrected). Working memory, letter and category fluency were equally impaired in FLE and TLE. Patients with FLE performed the verbal working memory task less well than controls, but similarly to those with TLE (>80% median accuracy in both patient groups). For visual working memory, performance in FLE was worse than controls during both task conditions, with more marked effects for higher task difficulty; there were no differences between FLE and TLE.

### Cognitive fMRI: synopsis

During language tasks, we found reduced activation of frontal areas and reduced deactivation of DMN nodes in FLE compared to controls. Abnormalities during working memory in FLE included reduced frontoparietal activation, reduced DMN deactivation, and global disorganization of task-related recruitment. For visual working memory, we observed a combination of (i) increased frontoparietal activation and lesser DMN deactivation than controls for low-level task demands, followed by (ii) reduced frontoparietal activation for higher task demands. Patterns of dysfunction in FLE broadly overlapped with those observed in TLE; DMN abnormalities, however, more evident in FLE, while reduced activation of posterior language areas was more marked in TLE.

The following sections present these findings in detail. For voxel-based analyses, the figures show regionally unconstrained whole-brain maps, and outline corrected as well as uncorrected findings for completeness, and in accord with benchmark evidence.^64^ As detailed above, however, voxel-based statistical tests focused on effects in prespecified cortical regions, and we only discuss findings that survive voxel-wise FWE-correction for multiple comparisons.

### Verbal fluency fMRI

In controls, the verbal fluency task activated fronto-temporo-parietal cortices, along with hippocampus and subcortical regions (Figure 1); deactivation encompassed DMN areas, including medial prefrontal, medial parietal and angular cortices. Analysis of systems provided an ensemble perspective on these findings, showing activation of cognitive control and salience systems (β= 0.10/0.08, *p*_FDR_=0.004/0.020), and tendencies for deactivation of the whole DMN at uncorrected thresholds (β=−0.06, *p*_unc_=0.038; see Supplementary Results for analyses across DMN subsystems). Gradient-based profiling sorted cortical regions according to a sensory-to-transmodal hierarchy, and showed: (i) positive task effects at the unimodal gradient end, reflecting activation of visual/primary sensory areas; (ii) positive effects along intermediate and right-sided segments, supporting the engagement of attentional (perceptually-coupled) and high-order executive processing; and (iii) a negative deflection at the transmodal gradient apex, capturing default-mode deactivation (all *p*_FDR_<0.05).

Compared to controls (Figure 2, Supplementary Table 1), FLE patients had reduced activation of left middle and inferior frontal gyrus, middle-anterior and middle-posterior temporal areas, (*p*_FWE_<0.05), and reduced deactivation of bilateral anterior and posterior DMN, left posterior temporal and angular gyrus (*p*_FWE_<0.05). FLE patients had similar cortical activation to the TLE group, but lesser deactivation of posterior temporal and anterior DMN areas (*p*_FWE_<0.05). Analysis of systems identified lower left-hemispheric cognitive control and limbic activity in FLE than controls (*p*_unc_=0.037/0.010; *d*=-0.30/-0.36), but no differences across whole-brain systems. DMN and cognitive control subdivisions undergoing deactivation in healthy controls showed lesser deactivation in FLE (DMN-A/DMN-C/control-C: *p*_FDR_=0.004/0.0025/<0.001, *d=*0.51/0.39/0.65; Supplementary Results, Supplementary Fig. 3). Curves of gradient-based task effects showed localized differences between FLE patients and controls, including (i) weaker task activity in intermediate gradient segments, and (ii) an increase at the transmodal apex, which implies lesser deactivation (all *p*_FDR_<0.05 across bins; *d=*-0.39 to -0.42 for intermediate bins, *d=*0.54 to 0.64 for bins at the apex). There were no differences between FLE and TLE for analyses of systems and gradients.

**Figure 2.**
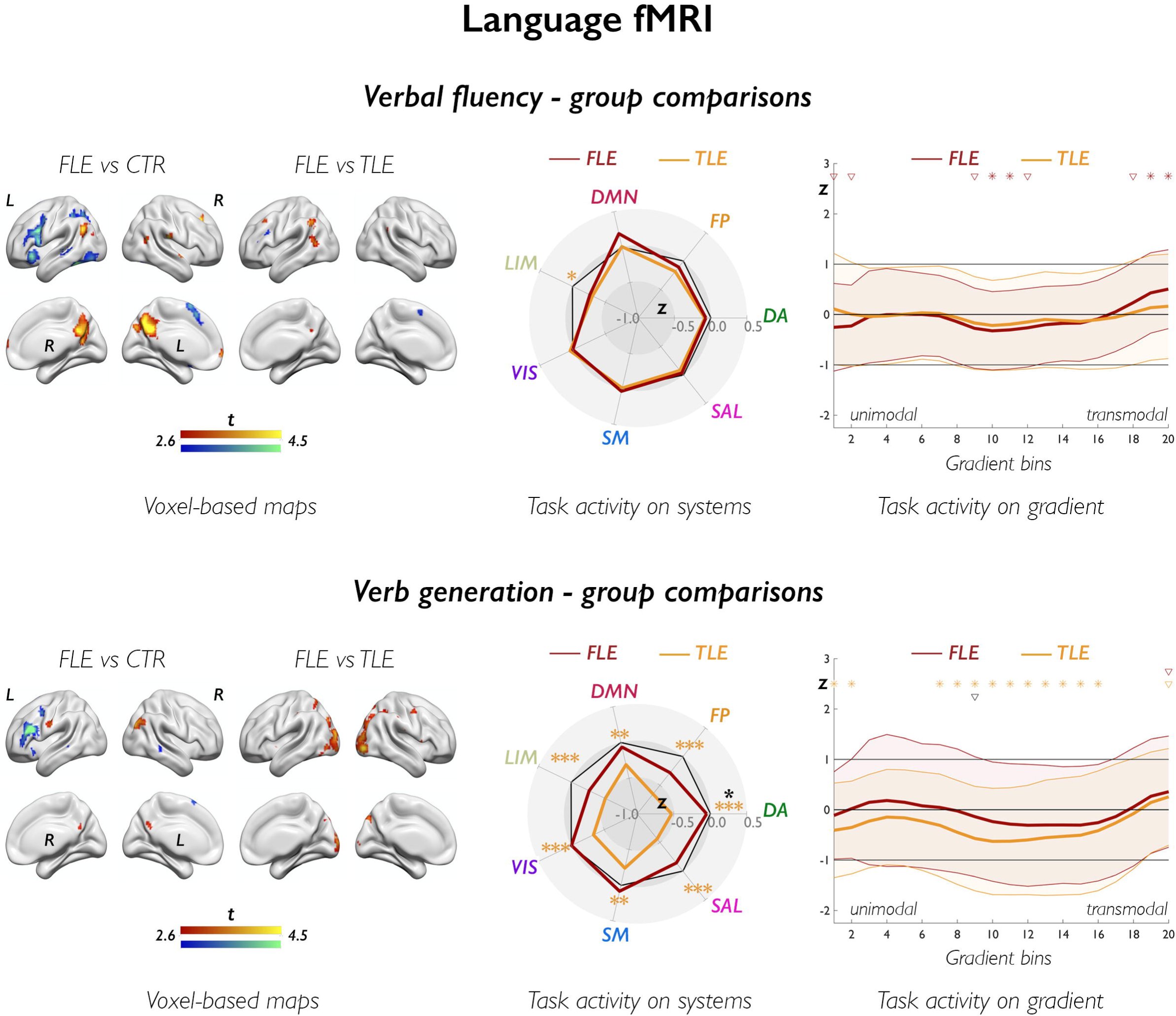
Language fMRI: group comparisons. The figure shows group comparisons for verbal fluency fMRI (upper panels) and verb generation fMRI (lower panels). In brain renders for comparison of patients with FLE and controls, cold and warm colors refer to lower and higher task-related effects in patients; of note, increases in FLE exclusively mapped on areas undergoing task-related deactivation, and are thus to be interpreted as areas of impaired deactivation in FLE vs controls; for comparison of FLE and TLE, cold and warm color scales refer to lower and higher task-related effects in FLE relative to TLE. Group differences are shown at p<0.005, with an extent threshold of 10 voxels applied for display purposes; color bars indicate corresponding t-score scales, MNI coordinates and statistical details for differences within prespecified regions of interest are provided in the Supplementary Material. The spider plots (middle panels) show Z-score analyses of task effects across 7 canonical systems (abbreviated as in Figure 1); across panels, black heptagons display effects in controls (Z-score =0, for each system), while effects in FLE and TLE are shown in dark red and orange lines and correspondingly colored asterisks, respectively; the dark asterisk in the verb generation spider plot highlights a difference between FLE and TLE; ***, *p*_FDR_<0.01; **, *p*_FDR_<0.05; *, uncorrected *p*<0.05. The upper and lower right panels show task-related effects, plotted as Z-scores (*y* axis), stratified along the principal gradient, which was discretized into 20 consecutive, equally-sized bins (*x* axis). Effects in FLE and TLE are shown in dark red and orange lines and in correspondingly colored asterisks, respectively; in each group, shaded areas correspond to one standard deviation; black asterisks refer to direct comparison of FLE and TLE; for each symbol, irrespective of color: *, *p*_FDR_<0.05; ∇, uncorrected *p*<0.05.

### Verb generation fMRI

In controls, the verb generation task activated fronto-temporo-parietal cortices and subcortical areas (Figure 1). Lateral temporal effects were more marked than in verbal fluency; moreover, the left posterior temporal cortex and angular gyrus belonged to the task activation map. Task activation involved cognitive control, DMN, salience and dorsal attention systems (β=0.12/0.06/0.08/0.06, *p*_FDR_<0.0001/0.017/0.0001/0.017, respectively). Gradient profiles indicated extensive activation across the intermediate-to-transmodal gradient segments (all *p*_FDR_<0.05).

Compared to controls (Figure 2, Supplementary Table 2), FLE exhibited lower inferior frontal activation and reduced right angular deactivation (*p*_FWE_<0.05). Compared to TLE, FLE had higher left posterior temporo-parietal and bilateral occipital activation, and reduced deactivation of the right angular gyrus and bilateral precuneus (all *p*_FWE_<0.05). Analysis of systems showed no corrected differences between FLE and controls, and between FLE and TLE; at uncorrected thresholds, there was lower left-hemispheric cognitive control activity in FLE than controls (*p*_unc_=0.011; *d*=-0.35), and better recruitment of the dorsal attention system in FLE than TLE (*p*_unc_=0.031, *d=*0.40). In FLE, comparison of gradient curves showed one uncorrected positive deviation from controls at the transmodal apex (*p*_unc_=0.017, *d=*0.32). There was one uncorrected difference between FLE and TLE at an intermediate gradient bin, with higher task activity in FLE (*p*_unc_*=*0.048, *d=*0.39).

### Verbal working memory fMRI

In controls, the verbal working memory task elicited bilateral frontoparietal activation, mapping to dorsal attention and control systems (β=0.20/0.26, *p*_FDR_*<*0.0001; Figure 3, Supplementary Fig. 4). Deactivation involved posterior cingulate cortex/precuneus, medial prefrontal cortex, and sensorimotor cortices (β=−0.08, *p*_FDR_*=*0.002, somatomotor effects). Gradient profiling showed positive shifts along the intermediate-to-transmodal segments, implicating attentional and executive processing, and decreases at the DMN apex (all *p*_FDR_<0.05).

**Figure 3.**
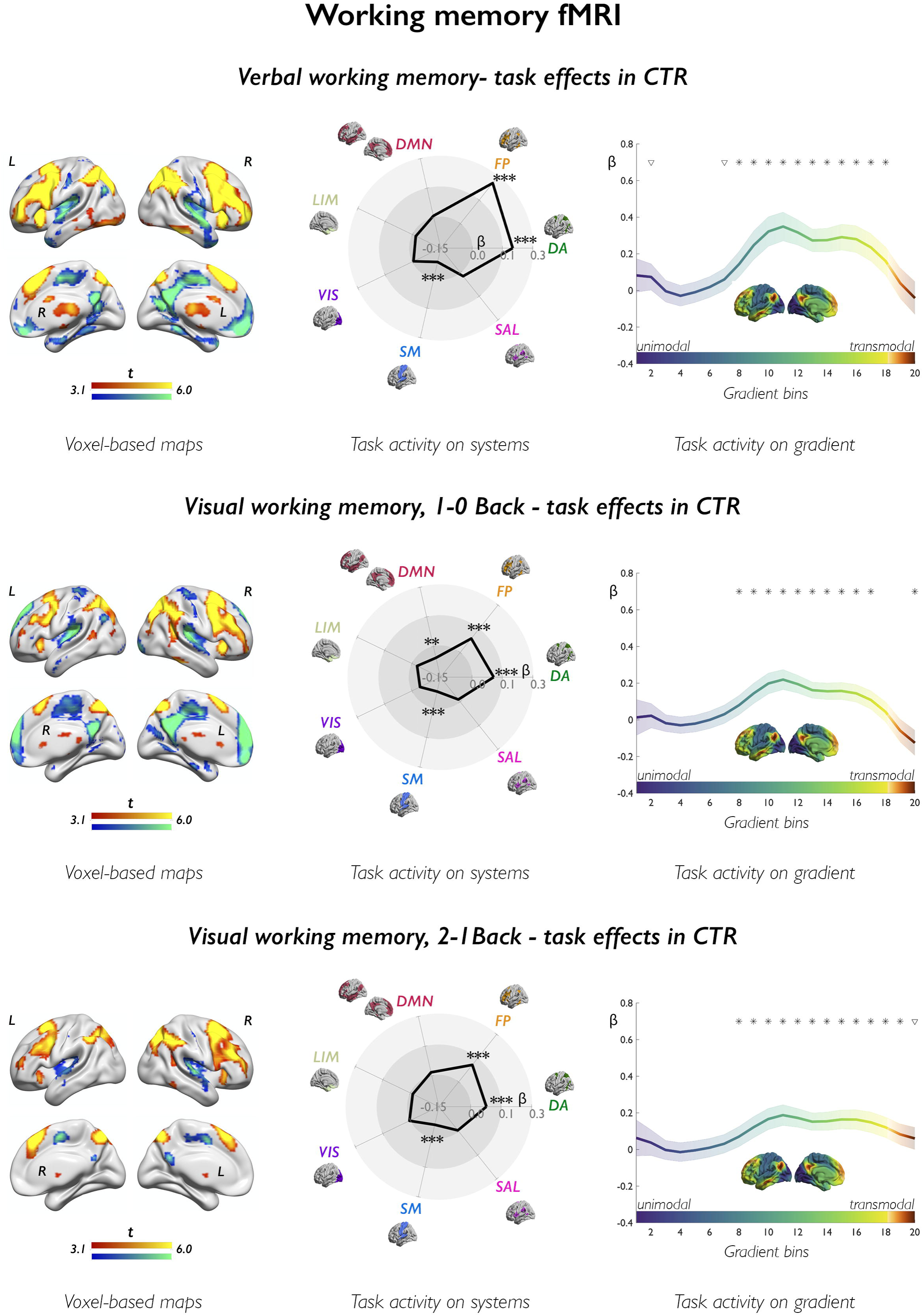
Working memory fMRI: task effects in controls. Across analytical scales, the upper, middle, and lower figure panels show task effects in controls for verbal working, 1–0Back visuo-spatial, and 2–1Back visuo-spatial working memory fMRI, respectively. Panels on the left show task-related activation (warm colors) and deactivation (cold colors) in controls, as derived from one-sample *t*-tests. Brain renders show maps at p<0.001, with an extent threshold of 10 voxels applied for display purposes; color bars indicate t-score scales. The spider plots (middle panels) show mean task-related effects, parameterized as β weights (contrast estimates) across 7 functional systems, where DA: dorsal attention; FP: cognitive (frontoparietal) control; DMN: default-mode network; LIM: limbic; VIS: visual; SM: somatomotor; SAL: salience (ventral attention); ***, *p*_FDR_<0.01; **, *p*_FDR_<0.05; *, uncorrected *p*<0.05. The upper and lower right panels show task-related effects stratified along the principal gradient; its left lateral and midline views are provided in the middle of each plot, using the same color scale as in curves of gradient-stratified task effects. The gradient was discretized into 20 consecutive, equally-sized bins (*x* axis), with task-related signal (β weights; *y* axis) computed per bin; shaded areas refer to 95% confidence intervals of mean effects at each bin; *, *p*_FDR_<0.05; ∇, uncorrected *p*<0.05.

Compared to controls, FLE patients had less frontoparietal activation and reduced deactivation of DMN areas (*p*_FWE_<0.05, Figure 4; Supplementary Table 3). Compared to patients with TLE, those with FLE showed reduced deactivation of posterior DMN areas (*p*_FWE_*<*0.05). Analysis of systems showed reductions of dorsal attention and cognitive control system activity in FLE than controls (*p*_FDR_<0.001/0.019, *d*=-0.62/-0.40). Gradient-stratified task profiles showed localized differences between FLE and controls along intermediate gradient segments (*p*_FDR_<0.05; *d* range=-0.52 to -0.33). There were no differences between FLE and TLE for analysis of systems and gradients.

**Figure 4.**
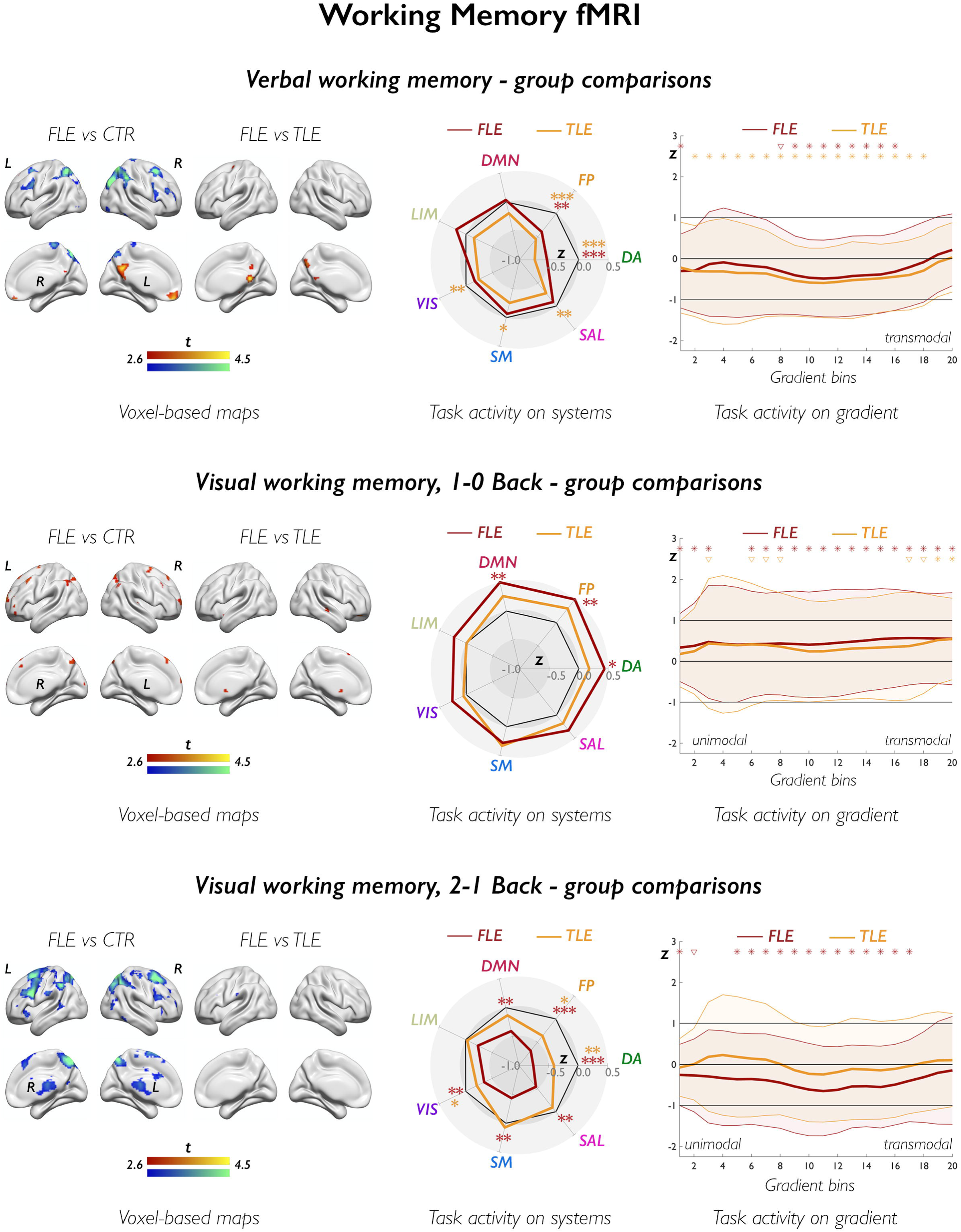
Working memory fMRI: group comparisons. The upper, middle, and lower figure panels show group comparisons for verbal, 1–0Back visuo-spatial, and 2–1Back visuo-spatial working memory fMRI, respectively. In brain renders (left-sided panels) for comparison of patients with FLE and controls, cold and warm colors refer to lower and higher task-related effects in patients; for comparison of FLE and TLE, cold and warm color scales refer to lower and higher task-related effects in FLE than in TLE. Group differences are shown at p<0.005, with an extent threshold of 10 voxels applied for display purposes; color bars indicate corresponding t-score scales, MNI coordinates and statistical details for differences within prespecified regions of interest are provided in the Supplementary Material. The spider plots (*middle panels*) show Z-score analyses of task-related signal across 7 canonical networks (abbreviations as in Figure 3); across panels, black heptagons display effects in controls (Z-score=0, at each system), while effects in FLE and TLE are shown in dark red and orange lines and in correspondingly colored asterisks, respectively; ***, *p*_FDR_<0.01; **, *p*_FDR_<0.05; *, uncorrected *p*<0.05. The z-score for cognitive control task activity in FLE, 1-0Back visual working memory, is 0.62; for display purposes, however, the spider plot axes reach a maximum of Z=0.5. The upper and lower *right* panels show task-related effects, plotted as Z-scores (*y* axis), stratified along the principal gradient, which was discretized into 20 consecutive, equally-sized bins (*x* axis). Effects in FLE and TLE are shown in dark red and orange lines and in correspondingly colored asterisks, respectively; in each group, shaded areas correspond to one standard deviation; black asterisks refer to direct comparison of FLE and TLE; for each symbol, irrespective of color: *, *p*_FDR_<0.05; ∇, uncorrected *p*<0.05. Statistical details are provided in the main text.

### Visual working memory fMRI

In controls, the 1–0Back contrast elicited bilateral frontoparietal activation and deactivation of midline DMN areas. Contrasting high versus low working memory demand (2–1Back) showed increasing recruitment of frontoparietal regions (Figure 3). Analysis of systems identified dorsal attention and cognitive control system activation (β=0.11/0.09, *p*_FDR_*<*0.0001/0.0002, 1– 0Back; β=0.08/0.11, *p*_FDR_=0.005/<0.0001, 2–1Back), DMN deactivation for the 1-0Back contrast (β=−0.05, *p*_FDR_=0.023), and somatomotor deactivation for both contrasts (β=−0.08 and −0.06, *p*_FDR_*<*0.0001 and 0.005, 1–0Back and 2–1Back). Gradient analyses indicated positive activity shifts along its intermediate to transmodal segments, and a significant decrease in correspondence of the default-mode apex (all *p*_FDR_<0.05 for both conditions).

For the 1–0Back contrast, the FLE group had increased parietal and dorsolateral frontal activation, and reduced deactivation of anterior DMN areas compared to controls (*p*_FWE_<0.05; Figure 4, Supplementary Table 4). Analysis of systems showed higher cognitive control and DMN activity in FLE than controls (*p*_FDR_=0.015/0.026, *d*=0.47/0.41), and increased dorsal attention activity at uncorrected thresholds (*p*_unc_=0.027, *d=*0.34). Gradient profiles globally differed in FLE compared to controls (FDA, *p*=0.022), with bin-wise analyses showing stronger activation in FLE across most gradient sections (*p*_FDR_<0.05, *d* range=0.31 to 0.51). There were no significant differences between FLE and TLE for voxel-based, system or gradient analyses.

For the 2–1Back contrast, FLE showed less frontoparietal activation than controls (*p*_FWE_<0.05; Figure 4; Supplementary Table 5). Across most systems, FLE had pronounced negative deviations from healthy controls, particularly for dorsal attention and cognitive control systems (*p*_FDR_<0.0001/0.007, *d=*-0.68/-0.56). Gradient-based profiles showed global disorganization of task-related recruitment in FLE compared to controls (FDA, *p=*0.034), with widespread involvement of intermediate and transmodal gradient segments (all *p*_FDR_<0.05; *d* range =-0.60 to -0.32). There were no significant differences between FLE and TLE for voxel-based, system or gradient analyses.

### Correlation of fMRI measures with cognitive performance

Across all domains, we identified significant correlations between cognitive scores and functional imaging metrics. All correlation analyses are detailed in the Supplementary Results.

For language fMRI, stronger frontal activation was associated with both higher verbal fluency and naming scores; naming also positively correlated with lateral temporal activation, particularly for verb generation fMRI. Conversely, less deactivation of bilateral precuneus during verbal fluency and right posterior temporal areas during verb generation related to worse fluency and naming performance, respectively (Figure 5; *p*_FWE_<0.05). Effect across systems were limited (*r*perm=-0.16, *p*_unc_*=*0.046, verb generation/naming correlation for the limbic system). For verbal fluency fMRI, gradient-based effects at the transmodal apex were negatively correlated with letter fluency (*r*perm=-0.17/-0.19, *p*=0.036/0.026 for significant bins).

**Figure 5.**
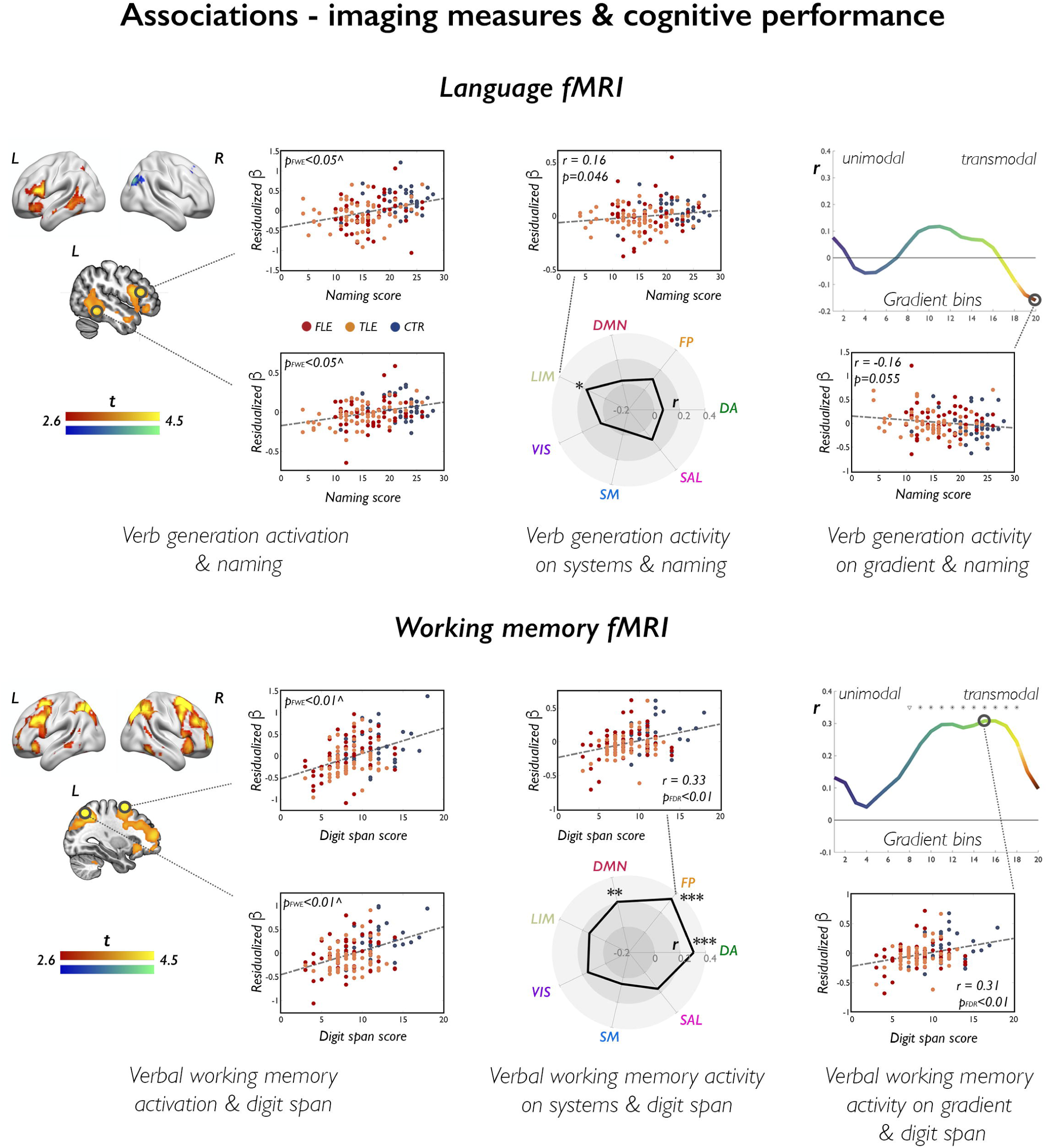
Correlations of functional imaging measures with cognitive performance. The brain renders and sections on the left display statistical maps of nonparametric multiple regression analyses probing associations between language fMRI (verb generation) and naming scores (upper panel), and between working memory fMRI (verbal task) and digit span scores (lower panels). Cold/warm color scales refer to negative/positive associations, respectively. Maps are shown at p<0.005, with an extent threshold of 10 voxels applied for display purposes; color bars indicate corresponding t-score scales. ^ Scatterplots highlight data distribution for the peak voxel within areas highlighted with a black circle; for illustration purposes, we used age- and sex-adjusted (residualized) parameter estimates (β) as measures of task effect. MNI coordinates and exact *p*-values are provided in the Supplementary Material. The spider plots (middle panels) and gradient plots (right-sided panels) show correlation coefficients for associations between cognitive measures (naming/digit span) and task effects (verb generation/verbal working memory) across each canonical system or each gradient bin. Example scatterplots are provided to highlight data distribution for correlations at the level of one given system or one given bin; for analyses of systems: ***, *p*_FDR_<0.01; **, *p*_FDR_<0.05; *, uncorrected *p*<0.05; for analyses along the gradient: *, *p*_FDR_<0.05; ∇, uncorrected *p*<0.05.

**Figure 6.**
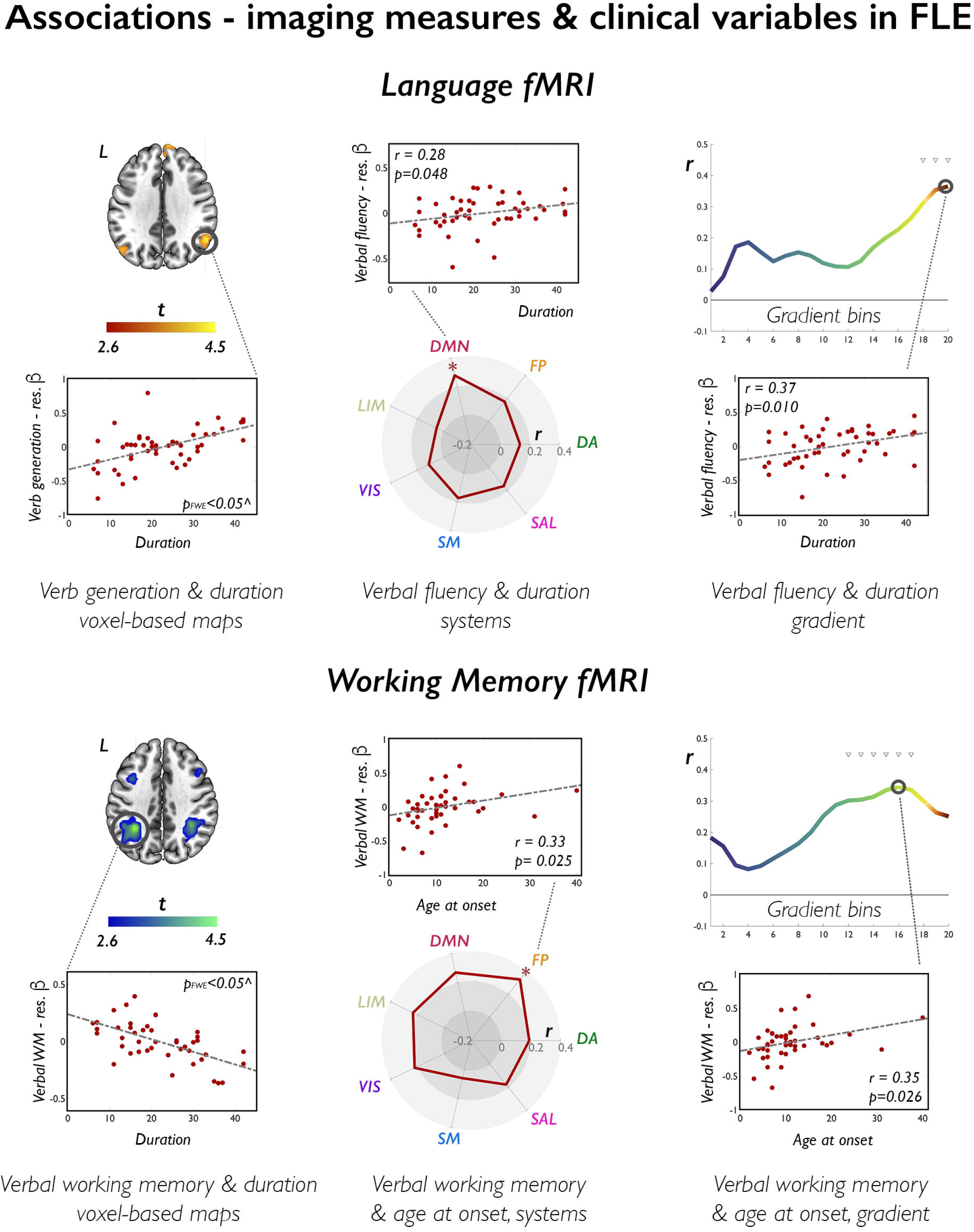
Correlations of functional imaging measures with clinical variables in FLE. The brain sections on the left show statistical maps of nonparametric multiple regressions probing associations between language or working memory fMRI and epilepsy duration in FLE. For language (upper panels), we show positive associations between duration of epilepsy and task effects during verb generation, where significant voxels (right posterior temporal/angular ROI, *p*_FWE_<0.05 within ROI; highlighted in black circle) map on areas undergoing task related deactivation; a positive association (warm color) indicates reduced deactivation with longer duration of disease. For working memory (lower panels), we show areas of negative associations between frontoparietal activation and disease duration with cold colors. Maps are shown at p<0.005, with an extent threshold of 10 voxels applied for display purposes; color bars indicate t-score scales. ^ Scatterplots highlight data distribution for the peak voxel within areas highlighted with a black circle; for illustration purposes, we used age- and sex-adjusted (residualized) parameter estimates (β) as measures of task effect. MNI coordinates and exact *p*-values are provided in the Supplementary Material. The spider plots (middle panels) and gradient plots (right-sided panels) show correlation coefficients for associations between epilepsy duration and task effects for verbal fluency fMRI, and between age at onset and verbal working memory fMRI, across each canonical system or gradient bin. Example scatterplots are provided to highlight data distribution for correlations at the level of one given system or one given bin; for analyses of systems: ***, *p*_FDR_<0.01; **, *p*_FDR_<0.05; *, uncorrected *p*<0.05; for analyses along the gradient *, *p*_FDR_<0.05; ∇, uncorrected *p*<0.05.

For working memory fMRI, digit span/visual 2Back performance scores strongly correlated with: (i) bilateral frontoparietal activation during verbal/2–1Back visual working memory at the voxel level (Figure 5; *p*_FWE_<0.05); (ii) activity of dorsal attention and cognitive control systems (*rperm*=0.31/0.33, *p*_FDR_=0.0007, for verbal working memory fMRI and digit span; *ρperm*=0.37/0.39, *p*_FDR_=<0.0001/0.0001, for 2–1Back fMRI and visual 2Back performance); (iii) task signal across intermediate and transmodal gradient sections (p_*FDR*_<0.05, permuted correlation ranges: 0.20 to 0.36). Similar patterns were evidenced for correlations between verbal 2Back task performance and verbal working memory fMRI (Supplementary Results).

### Correlation of fMRI measures with clinical variables

For language tasks in FLE, longer epilepsy duration related to: (i) stronger verbal fluency fMRI activity in a rostral left middle frontal area bordering the classical activation map (*p*_FWE_<0.05), possibly reflecting compensatory recruitment; (ii) lesser deactivation of the DMN as a whole (*r*perm= 0.28, *p*_unc_*=*0.048), and (iii) lesser deactivation at the gradient apex (bins 18-20, *r*perm range*=*0.31 to 0.37, *p*_unc_*=*0.010 to 0.029); longer duration was also associated with lesser posterior-temporal and angular deactivation during verb generation (*p*_FWE_<0.05). For working memory in FLE, we found a significant association between lower bilateral parietal activation during the verbal task and longer disease duration (*p*_FWE_<0.05). This was paralleled by positive correlations between age at onset and (i) cognitive control activity (*r*perm*=*0.33, *p*_unc_*=*0.025), as well as (ii) gradient-based profiles (bins 12-18, *r*perm range*=*0.30 to 0.35, *p*_unc_*=*0.026 to 0.048) during verbal working memory. In TLE, earlier age at onset was associated with lower inferior frontal activation during verbal fluency, and longer duration related to lesser deactivation of posterior DMN areas during working memory at the voxel level (*p*_FWE_<0.05). Further statistical details are available in the Supplementary Results.

### Sensitivity analyses

*a) Subgroup analysis in FLE with FCD*: We separately investigated an FLE subgroup with suspected focal cortical dysplasia (FLE-FCD; n=13, pathologically confirmed in 8). Across domains and analysis scales, comparisons of the FLE-FCD subgroup to controls produced results similar to those in the main group comparisons (Supplementary Results, Supplementary Fig. 5).
*b) Mitigation of the effects of frontal language laterality:* Analysis of language laterality indices (LIs) showed weaker left lateralization of frontal hemispheric dominance for language in FLE compared controls (Table 1). Repeat voxel-based analyses covarying for language LI in frontal regions of interest, however, still yielded significant left frontal activation differences between FLE and controls for both verbal fluency and verb generation tasks (*p*_FWE_<0.05; Supplementary Table 6).
*c) Separate analysis of left and right FLE:* Lateralization of the seizure focus may differentially affect cognitive system activity, particularly for verbal tasks. We thus conducted repeat voxel-level analyses separately comparing left and right FLE subgroups to controls. For language tasks, left and right FLE had similar differences compared to controls (Supplementary Fig. 6). Patterns in left and right FLE were also similar for the verbal working memory task. For visual working memory, there were more marked frontoparietal activation and impaired frontal DMN deactivation in left FLE, for the 1-0 Back contrast, and more marked activation decreases in right FLE for the 2-1Back contrast (Supplementary Fig. 6).

## DISCUSSION

Knowledge of the neurobiological substrates of cognitive impairment in FLE is scarce. Here, we profiled the neural correlates of language and working memory dysfunction in a large FLE sample. Concomitant analysis of a TLE group allowed us to decode shared and syndrome-specific effects. Our findings indicate impaired fronto-temporo-parietal activation and impaired DMN deactivation in FLE, and implicate areas across a broad spectrum of functional specialization. Global disorganization of task-related recruitment, as elucidated by system and gradient analyses, was evident for working memory. While patterns of dysfunction largely overlapped across syndromes, we found more prominent alterations of DMN deactivation in FLE, and more marked temporal abnormalities in TLE during verb generation, which entails semantic processing. Task profiles in FLE were detrimentally modulated by clinical characteristics. This study conveys a comprehensive characterization of the neural correlates of cognitive impairment in FLE, paving the way for future analyses of disease subgroups. We provide neural targets that may aid future investigations into cognitive prognostics, and inform the development of novel rehabilitation and therapeutic approaches.

Neuropsychological profiles indicated generalized cognitive impairment in FLE, with poorer performance for functions typically ascribed to the frontal lobes, including working memory, verbal fluency and mental flexibility, and weaker verbal memory and naming, that rely more markedly on temporal lobe processing. Comparison of FLE and TLE showed more prominent executive dysfunction in the former, and more marked impairment of verbal learning and semantic knowledge in the latter. Our findings corroborate evidence of dysexecutive traits and memory difficulties in FLE,^9, 10, 12^ and indicate that cognitive profiles in FLE and TLE differ. Echoing prior work,^4, 8, 15^ we suggest that functions that more strongly recruit areas overlapping with the epileptic network are affected more pervasively.

Traditional fMRI activation maps provide a fine-grained account of regional group differences, but do not capture large-scale effects. With our multiscale approach, we also attained systems-level inference^36, 62, 65^ and profiled cognition on the backbone of an organizational principle of intrinsic connectivity, the principal gradient.^37, 66^ The gradient describes a continuous transition from unimodal sensory to transmodal areas, and offers a compact framework to conceptualize task-related effects in the context of cognitive system hierarchies.^40, 41, 57^ Our analyses enabled decoding of the neural signatures of cognition from a “local-to-global” perspective, captured the landscape of competing activation and deactivation during cognition, and provide a novel framework to investigate cognitive impairment in epilepsy.

In controls, language tasks elicited left-lateralized activation of middle and inferior frontal gyrus and temporo-parietal cortices, which positively correlated with fluency and naming performance. Systems-level effects captured cognitive control and opercular involvement, and activity shifts along the intermediate to transmodal gradient segments implicated a combination of attentional and high-level executive processing. Suppression of DMN activity was extensive for verbal fluency, which requires executive control,^16, 67^ was tracked by negative effects at the transmodal gradient apex, and was neurobehaviorally relevant, as demonstrated by correlations analyses with out-of-scanner performance. Temporo-parietal activation was more marked during verb generation, in light of its semantic demands,^68, 69^ and correlated with naming.

Across language tasks, we found reduced activation of the left inferior and middle frontal gyrus in FLE, which represented an isolated abnormality during verb generation, but coexisted with temporal and subcortical differences during verbal fluency. We also observed slightly weaker frontal language lateralization in FLE compared to controls, corroborating findings of prior case series.^70, 71^ Repeat group comparisons controlling for frontal language laterality, however, did not change our main results, excluding a substantial influence of interhemispheric frontal language organization on the observed group differences. Thus, dysfunction of the left frontal language core may be a key feature underlying impaired expressive language in FLE. From a neurobiological perspective, such dysfunction may be a result of epileptic activity of frontal origin, with adverse effects on synaptic connections and local computation. Exploratory analyses identified similar disruptions of left-hemispheric activation in left and right FLE. Involvement of left-hemispheric language areas and verbal deficits were previously described in people with right TLE, which led to reconsider earlier views on the sparing of verbal functions in right-hemisphere pathology.^16, 72–74^ Along the same lines, it is possible that left frontal language alterations in right FLE may be a consequence of transcallosal propagation of epileptic activity,^76^ given the rapid bilateral spread of frontal lobe seizures.^1, 75^

FLE and TLE groups had similar frontal abnormalities during verbal fluency. Frontal language dysfunction thus represents a shared trait, possibly as a downstream consequence of both proximal (frontal) and more distal (temporal) pathology. Notably, we and others previously showed abnormal frontal activation and frontotemporal connectivity during expressive language fMRI in TLE.^22, 23, 77, 78^ Alteration of frontal function in TLE may emerge as propagated abnormality, and could be mediated by microstructural alterations in perisylvian white matter tracts.^49, 73, 74^ During verb generation, there were differences between FLE and TLE in the semantic processing stream, involving posterior temporo-parietal and occipital areas.^69, 79, 80^ More marked impairment during tasks with semantic demands thus appears TLE-specific, and correlates with neuropsychological findings. While frontal language areas represent a common substrate of expressive language dysfunction across TLE and FLE, temporal pathology may thus primarily affect areas in its vicinity, such as posterior language centers, leading to more pervasive dysfunction during tasks that specifically rely on these.

In both working memory tasks, we detected bilateral frontoparietal activation,^31, 81^ that mapped on dorsal attention and cognitive control systems, resulted in activity shifts in the intermediate to transmodal gradient sections, and strongly correlated with in-scanner task performance and digit span scores. For both verbal and visual conditions entailing a 2–Back working memory span, FLE exhibited bilateral attenuation of frontoparietal activation and globally altered gradient profiles, which occurred in both left and right FLE. Activation reductions were particularly marked for the 2–1Back contrast, and comprised cognitive systems beyond those directly implicated in working memory. While indicating global dysfunction, such widespread effects may also reflect loss of motivation and participant disengagement. The verbal working memory task, however, was less challenging and, though performed slightly less accurately by people with FLE than controls, resulted in satisfactory performance (>80%) in all groups. Patients with FLE still presented with frontoparietal hypoactivation, more selective for dorsal attention and cognitive control systems. Thus, we suggest that attenuated frontoparietal activation may more parsimoniously reflect inefficient recruitment of areas required for successful task performance. The low-demand visual working memory contrast, on the other hand, was associated with enhanced frontoparietal activation and reduced DMN deactivation in FLE. This sequence of higher and lower activation for easy and difficult task conditions points to cognitive system saturation that already occurs for low-level task demands, and is followed by defective additional recruitment for higher task difficulty levels. Notably, by encompassing altered deactivation of DMN regions, neural processes underlying working memory in FLE already proved inefficient for easier task conditions.

Comparison of FLE and TLE for working memory did not indicate marked group differences. Working memory relies on the activity of distributed, bilateral fronto-temporo-parietal networks.^35, 82, 83^ It is possible that propagation of ictal and interictal epileptic activity, and consequent long-lasting neural derangements within multiple sites relevant for performance, all result in globally less efficient working memory networks, independent of the location of the area of seizure onset. Future investigation of recent-onset focal epilepsy may clarify whether the involvement of working memory hubs may be sequential, with earlier effects close to the seizure focus.

Across tasks, we showed impaired deactivation of DMN areas in FLE, and to a lesser extent in TLE. The DMN subserves processes including self-awareness, mind-wandering, and cognition supported by internal representations.^84–86^ DMN deactivation and anti-correlation between activity patterns in DMN regions and frontoparietal areas were described for tasks with executive demands.^33, 87, 88^ Alterations in such processes were previously identified in a spectrum of psychiatric disorders, including autism^89, 90^ and schizophrenia,^91, 92^ and may reflect suboptimal distribution of neural resources during goal-directed cognition.^93^ We and others previously showed altered DMN deactivation during working memory in JME and pediatric TLE.^47, 94^ Our current findings in FLE underscore the vulnerability of the DMN across the epilepsy spectrum, and indicate altered DMN deactivation profiles as a possible trans-syndromic marker. We previously documented an influence of ASMs on task-related deactivation during language and working memory.^44, 45, 95^ Such effects, however, largely spared midline anterior and posterior DMN areas, which, in this study, were associated with prominent differences between FLE and controls, and between FLE and TLE. Interestingly, alterations of task-related deactivation, mostly encompassing DMN nodes, were more marked in FLE than TLE. Future connectivity-based analyses may better characterize the underlying sources of such phenomenon; a potential explanation may lie in the differential connectivity profiles of specific DMN hubs, such as the medial prefrontal cortex, that is strongly embedded within midline DMN, and likely to be involved in the propagation network of frontal seizures.^75^

Correlation analyses showed that disease load, as tracked by age at onset and epilepsy duration, may modulate severity of both activation and deactivation profiles in FLE. Such effects were particularly evident for verbal working memory. Prior work has identified detrimental associations between cognitive function and early age at epilepsy onset,^96–99^ that are possibly linked to disrupted white matter maturation.^100, 101^ In our study, associations were more evident for functions that have a prolonged maturational trajectory, such as working memory, which depends on the myelination of long-range fiber tracts.^102, 103^

The co-occurrence of impaired frontoparietal activation and DMN deactivation in FLE mirrors findings of studies comparing healthy older adults (∼70 years) to younger individuals.^104, 105^ As the mean age in our FLE sample was ∼30 years, this raises the question of whether the identified traits may reflect a functional marker of accelerated brain aging. If proven by future work, such phenomenon would dovetail with recent evidence of accelerated grey matter loss in people with focal epilepsy, which revived the “disease progression” hypothesis.^106–109^

Our study has several strengths, including the use of large samples, robust methodology, comprehensive mapping of function from local to global perspectives, direct comparison of patient groups, and several sensitivity analyses. Our study also has limitations. The FLE group was heterogeneous in terms of etiology and MRI findings, but is representative of the spectrum of FLE patients assessed in tertiary centers.^3, 9, 110^ In addition, sensitivity analyses in the FLE-FCD patient subgroup largely corroborated our main findings. Future work is encouraged to disentangle cognitive alterations specific to distinct etiologies, and to patient subgroups based on sub-lobar lesion location or patterns of seizure semiology.^75^ The language tasks were covert, which prevented online performance monitoring. However, these tasks were previously validated,^49, 77^ are extensively used clinically,^111, 112^ and captured interindividual differences in out-of-scanner fluency and naming performance in our study. Patients with FLE and TLE were drug-resistant, taking ASMs in various combinations. We previously described compound- and syndrome-specific effects of ASMs on cognitive networks.^95, 113^ FLE and TLE groups were balanced for medications with known detrimental (topiramate/zonisamide) or more favorable (levetiracetam) cognitive network profiles.^44, 45^ As for comparisons with controls, it is possible that impaired task-related activation in FLE (and TLE) may be partially influenced by ASMs. As discussed earlier, however, previously described ASM-related effects on task deactivation patterns largely spared areas showing marked intergroup differences in this study. Finally, we provided evidence of associations between age at onset, disease duration and task-related network phenotypes, which suggests disease-related mediating factors other than ASMs. Future longitudinal work, including the assessment of drug-naïve patients, may better characterize the contribution of ASMs to cognitive system dysfunction in epilepsy.

## CONCLUSION

Our study maps language and working memory dysfunction in FLE, showing local, systems-level, and global abnormalities that indicate an altered interplay between task-related cognitive system activation and deactivation. While patterns of dysfunction in FLE and TLE largely overlap, activity of posterior language centers and profiles of default-mode deactivation are more and less favorably affected in FLE than TLE. This work bridges a substantial knowledge gap in the epilepsy literature, and delivers neural markers that can be validated in the context of cognitive prognostics, and may serve as targets for future development of targeted treatment.

## CITATION DIVERSITY STATEMENT

Recent work in several fields of science has identified a bias in citation practices such that papers from women and other minority scholars are under-cited relative to the number of such papers in the field.^114–118^ Here we sought to proactively consider choosing references that reflect the diversity of the field in thought, form of contribution, gender, race, ethnicity and other factors. First, we obtained the predicted gender of the first and last author of each reference by using databases that store the probability of a first name being carried by a woman^118, 119^. By this measure (and excluding self-citations to the first and last authors of our current paper), our references contain 10.71% woman(first)/woman(last), 10.71% man/woman, 27.38% woman/man, and 51.19% man/man. This method is limited in that a) names, pronouns and social media profiles used to construct the databases may not, in every case, be indicative of gender identity, and b) it cannot account for intersex, non-binary, or transgender people. Second, we obtained predicted racial/ethnic category of the first and last author of each reference by databases that store the probability of a first and last name being carried by an author of color.^120, 121^ By this measure (and excluding self-citations), our references contain 8.05% author of color (first)/author of color(last), 13.50% white author/author of color, 18.88% author of color/white author, and 59.57% white author/white author. This method is limited in that a) names, Census entries, and Wikipedia profiles used to make the predictions may not be indicative of racial/ethnic identity, and b) it cannot account for Indigenous and mixed-race authors, or those who may face differential biases due to the ambiguous racialization or ethnicization of their names. We look forward to future work that could help us to better understand how to support equitable practices in science.

## ABBREVIATIONS

ASM: anti-seizure medication
DMN: default-mode network
FCD: focal cortical dysplasia
FDA: functional data analysis
FDR: false discovery rate
FLE: frontal lobe epilepsy
fMRI: functional magnetic resonance imaging
FSL: FMRIB Software Library
FWE: familywise error (rate)
FWHM: full width at half-maximum
HCP: Human Connectome Project
LI: laterality index
MNI: Montreal Neurological Institute
NHNN: National Hospital for Neurology and Neurosurgery
ROI: region of interest
S(n)PM: Statistical (non-) Parametric Mapping
TLE: temporal lobe epilepsy.

## ACKNOWLEDGEMENTS

We thank our patients and controls for their participation in this study, and the radiographers at the Epilepsy Society MRI Unit, Philippa Bartlett, Jane Burdett, and Elaine Williams for their help during data acquisition. We also thank Jason Stretton for data acquisition. Dr. Jonathan O’Muircheartaigh and Professor Mark P. Richardson are acknowledged for previous collaborative research initiatives. Alexander Lowe is acknowledged for insightful discussions during prior work. Sara Larivière and Reinder Vos De Wael are acknowledged for helpful conversations on functional gradient analyses.

## SUPPLEMENTARY MATERIAL

Supplementary material is available for this manuscript.

