## Supplementary Material for "Disorganization of language and working memory systems in frontal versus temporal lobe epilepsy"


**Running title:** Mapping cognitive dysfunction in FLE versus TLE

**SUPPLEMENTARY METHODS** **4**
Additional clinical details 4
Neuropsychological tests 5
Functional MRI data: acquisition protocol, data pre-processing, quality checks 5
Structural data: acquisition protocol and surface-based pre-processing 6
Voxel-based fMRI, group comparisons: regions of interest and further statistical details 7
Supplementary Figure 1: Language, WM and task-negative ROIs 8
Voxel-based fMRI, language laterality indices 9
Curves of gradient-based task-effects: group comparisons via functional data analysis 9
**SUPPLEMENTARY RESULTS** **10**
Verbal fluency fMRI: comparison of TLE and controls 10
Verb generation fMRI: comparison of TLE and controls 10
Verbal WM fMRI: comparison of TLE and controls 10
Visual WM fMRI: comparison of TLE and controls 10
Supplementary Figure 2: Comparison of TLE and CTR, voxel-based analyses 12
Analyses across 17 canonical subsystems: description of system partition 13
Analyses across 17 canonical subsystems: verbal fluency fMRI (Supplementary Figure 3) 13
Analyses across 17 canonical subsystems: verb generation fMRI (Supplementary Figure 3) 14
Analyses across 17 canonical subsystems: verbal WM fMRI (Supplementary Figure 4) 14
Analyses across 17 canonical subsystems: visual WM fMRI (Supplementary Figure 4) 15
Supplementary Figure 3: analyses across 17 systems, language fMRI 17
Supplementary Figure 4: analyses across 17 systems, WM fMRI 18
Sensitivity analyses: FLE-FCD versus controls, language fMRI 19
Sensitivity analyses: FLE-FCD versus controls, WM fMRI 19
Supplementary Figure 5: Comparison of FLE-FCD and CTR, voxel-based analyses 21
Supplementary Figure 6: Analysis of left and right FLE subgroups 22
Correlations of fMRI measures and cognitive performance: language fMRI 23
Correlations of fMRI measures and cognitive performance: WM fMRI 24

Correlations of fMRI measures with age and onset and disease duration: FLE 24

Correlations of fMRI measures with age and onset and disease duration: TLE 25

**SUPPLEMENTARY TABLES 26**

Supplementary Table 1. Verbal fluency fMRI 26

Supplementary Table 2. Verb generation fMRI 27

Supplementary Table 3. Verbal WM fMRI 28

Supplementary Table 4. Visual WM fMRI, 1-0 Back 29

Supplementary Table 5. Visual WM fMRI, 2-1 Back 30

Supplementary Table 6. Language fMRI: frontal group differences covaried for language LI 31

Supplementary Table 7. Language fMRI and cognitive test scores 32

Supplementary Table 8. WM fMRI and cognitive test scores 33

**SUPPLEMENTARY REFERENCES 34**

### SUPPLEMENTARY METHODS

*Additional clinical details*

Main demographic and clinical details are provided in Table 1.

In patients with drug-resistant FLE, diagnosis was initially determined by expert epileptologists on the basis of history, seizure semiology, prolonged scalp video-EEG telemetry, and structural MRI at 3T; PET as well as ictal SPECT and/or magneto-encephalography data were also available for a patient subset. Patients were included in this analysis only if sufficient concordant information was available to determine lateralization of the epileptic focus. Seizure onset was lateralized to left frontal areas in 30 patients, and to right frontal areas in 26. In terms of etiological categories, there were no discernible lesions on MRI in 29 subjects (51.8%; 17 left, 12 right); in the remainder individuals, findings included areas of suspected focal cortical dysplasia (FCD; n=13, 23.1%, 6 left, 7 right; pathologically confirmed in 8 out of 8 patients who subsequently proceeded to surgery); dysembryoplastic neuroepithelial tumor (DNET; n=6, 10.7%; 3 left, 3 right); low-grade glial tumor (n=3, 5.4%, all right); one finding labelled as possible periventricular nodular heterotopia (n=1, 1.8%, left); or areas of unequivocal signal abnormalities concordant with clinical and EEG findings [n=4; 7.1%, three left and one right patients; one post-traumatic in nature, one of possibly intrauterine (vascular) etiology, and two representing clear areas of cortical injury of otherwise unclear etiology]. One incidental calcified cerebellar lesion of possible vascular etiology (cavernoma) was found in one patient, and nonspecific T2-FLAIR bifrontal hyperintensities were found in two other patients.

The TLE sample consisted of drug-resistant individuals under surgical consideration, who all underwent prolonged interictal and ictal scalp video-EEG, confirming and lateralizing seizure onset to the temporal lobe (34 left, 30 right). Patients with TLE had ipsilateral hippocampal sclerosis, as evidenced via 3T-MRI scans and qualitative diagnostic assessments conducted by experienced neuroradiologists, and/or via quantitative assessments of hippocampal volumes[e1] and T2 relaxation times,[e2,e3] and with pathological confirmation in those who subsequently proceeded to surgery. Hippocampal sclerosis co-existed with ipsilateral DNET in three patients (2 left, 1 right) and with an ipsilateral possible FCD in one (right).

*Neuropsychological tests*

General intellectual level (IQ) was assessed via the National Adult Reading Test.[e4] We probed working memory (WM) with the digit span subtest of the Wechsler Intelligence Scale (WAIS III);[e5] digit span raw scores were converted into scaled score equivalents, according to the WAIS III reference manual. Verbal fluency was tested with letter fluency (sum of words generated for letter "S" in one minute)[e6] and category fluency tests (sum of items generated for the category "Animals" in one minute).[e6] Visual confrontation naming was assessed with the McKenna Graded Naming Test).[e7] The Trail Making Test (TMT)[e8] assessed psychomotor speed (TMT A) and executive function (mental flexibility; TMT B-A). Verbal and visuo-spatial learning were tested with the List and Design Learning Subtasks of the Adult Memory and Information Processing Battery (Trials A1-A5; Trial A6, for delayed recall).[e9] Pairwise deletion was used in case of missing data.

In patient groups only, measures of verbal reasoning and comprehension [Vocabulary and Similarities subtasks, Wechsler Adult Intelligence Scale (WAIS III)][e5] were also available. For each patient group, vocabulary and similarities scores were converted into scaled score equivalents according to the WAIS III reference manual, and compared against published norms (mean=10) via one sample *t*-tests.

*Functional MRI data: acquisition protocol, data pre-processing, quality checks*

Imaging data were acquired on a GE Signa HDx 3T MRI scanner. For all tasks, we used a 50-slice gradient echo-planar sequence with axial orientation, 64x64 matrix, corresponding to in-plane voxel size of 3.75x3.75mm, 2.4mm slice thickness, 0.1mm inter-slice gap, echo time/repetition time: 25/2500ms.[47] Visuo-spatial/verbal WM fMRI data were available for 50/53 out of 56 patients with FLE, 62/63 out of 64 patients with TLE, and 52/51 out of 52 controls. Verbal fluency/verb generation fMRI data were available for all patients with FLE, 52/51 out of 52 controls, and 63/63 out of 64 patients with TLE. In SPM12, functional time series were realigned, resliced to 3x3x3mm, normalized to a scanner and acquisition-specific template in Montreal Neurological Institute (MNI) space,[47] and smoothed using a Gaussian kernel of 8x8x8mm full-width-at-half-maximum (FWHM), as previously.[51,e10] Excessive motion (mean framewise displacement >0.5mm)[52] was observed for 3/1/3/1 FLE, 5/4/4/3 TLE, and 2/2/1/2 control participants for visual WM/verbal WM/verbal fluency/verb generation fMRI, respectively. Participant maps were individually reviewed to rule out gross artefact, which led to the exclusion of visual WM fMRI data for one FLE and one TLE patient, and verbal WM fMRI data of one FLE patient. Some voxels within a frontal DNET were assigned a beta coefficient values of “not-a-number” during SPM-based model specification in one patient; these were operationalized as zeros for subsequent analysis passages. As language paradigms were covert, performance measures for in-scanner task execution were not available. As in previous work,[95,e10] individual-level activation of task relevant areas, such as fronto-temporal language regions, cerebellum, supplementary motor area and anterior insula, was assessed up to at an uncorrected threshold of $p<0.01$; lack thereof led to participant exclusion. We further excluded verbal working-memory fMRI data of one TLE patient, and verb generation fMRI data of one control owing to heavily corrupted field of view. Final analyses for visual WM/verbal WM/verbal fluency/verb generation fMRI included 46/51/53/55 FLE patients, 56/58/59/60 TLE patients, and 50/49/49/47 controls.

*Structural data: acquisition protocol and surface-based pre-processing*

Structural images were only used to align individual functional space to structural space before conversion of task maps to surface space in the context of gradient-based fMRI analyses. T1-weighted images were acquired on the same scanner as the functional images in all participant, with a coronal volumetric fast-spoiled-gradient-echo (FSPGR) sequence, matrix 256x256, slice thickness 1.1mm, in-plane resolution: 1.1x1.1mm. In patients with TLE and in a subset of controls (n=27), volumetric T1-weighted images had a slightly different in-plane resolution of 0.9375x0.9375mm; this was not associated with differences in MRI contrast between grey and white matter, and did not influence the quality of cortical mantle extraction. In each participant, we derived cortical surfaces using FreeSurfer (v6.0; http://surfer.nmr.mgh.harvard.edu/); processing pipelines in FreeSurfer have been detailed elsewhere,[e11,e12] and include reorientation, skull-stripping, tissue segmentation, generation of pial and white matter surfaces. T1-weighted data for one patient with FLE and one control were affected by heavy motion artefacts and were considered unusable for cortical surface extraction; these two subjects were thus discarded from gradient-based analyses in surface space.

For gradient-based stratification of functional data, task-based general linear models were computed in native space after SPM-based realignment for each subject, with task contrast specifics as detailed in the main manuscript text for all tasks; 6 motion parameters were included as nuisance regressors. Next, we used boundary-based registration[e13] to co-register cortical surfaces with mean functional and task contrast images. We then performed spherical mapping to the *fsaverage5* surface template (20484 vertices), and surface smoothing of the functional contrasts with a Gaussian filter of 5mm FWHM.

*Voxel-based fMRI, group comparisons: regions of interest and further statistical details*

For voxel-based analyses of language tasks, we assessed group differences within comprehensive frontal and temporo-parietal regions of interest (ROIs); to this end, we used previously validated ROIs,[23,e14,e15] that were defined functionally via a language-localizer fMRI paradigm in a sample of healthy participants,[e14] and included: (1) orbital inferior frontal gyrus, (2) inferior frontal gyrus, (3) middle frontal gyrus, (4) anterior and (5) middle-anterior temporal lobe, (6) middle-posterior and (7) posterior temporal lobe, as well as (8) angular gyrus. These ROIs were mirror-projected onto the right hemisphere to create homologous masks; for group comparisons, 8 bilateral ROIs were thus used.

For WM tasks, we implemented the following bilateral ROIs, based on prior meta-analytical work:[31,81] (1) one frontal-eye field/premotor ROI [Brodmann areas 8 and 6 (dorsal)]; (2) one dorsolateral/ventrolateral prefrontal ROI (Brodmann areas 9/46 and 44); and (3) one dorsal parietal ROI, encompassing dorsal precuneus, superior parietal lobule, part of the inferior parietal lobule, and rostral lateral superior occipital gyrus (Brodmann areas 7, 39 and 40). These three ROIs were obtained by merging the relevant Brodmann labels, provided via the *Brainnetome* Atlas.[e16]

For both tasks, we also assessed differences across midline default-mode (DMN) areas, belonging to the deactivation map, henceforth labeled as "task-negative" ROIs;[45,47,104,e17] to this end, we used : (1) one midline prefrontal ROI, that combines Brodmann areas 32 (anterior cingulate), 9/10 (rostral medial superior frontal), 11 and 14 (medial frontal/orbital); and (2) one midline parietal ROI, corresponding to the precuneus and posterior cingulate cortex (Brodmann areas 31 and 23). For language tasks, the midline parietal ROI additionally included the dorsomedial precuneus (part of Brodmann area 7), which has instead been described as part of the activation network for WM tasks,[31] and was therefore not included in the posterior DMN WM ROI. All ROIs are displayed in *Supplementary Figure 1*.

All ROIs were chosen *a priori*; comprehensive coverage of task-related activation/deactivation maps was qualitatively confirmed by overlapping those with unthresholded statistical maps of task-related effects derived from one-sample *t*-tests in controls. Of note, while frontal language ROIs belonged to the activation map in both language tasks, task-related effects within anterior, middle-anterior, middle-posterior temporal and angular ROIs could occur either in the form of activation or deactivation, depending on the specific task. Such regions are thus shown with superimposed dashed lines in *Supplementary Figure* 1. For each language task, intergroup differences within such ROIs are discussed in the form activation or deactivation differences on the basis of their task-related behavior in healthy controls.

For all voxel-based analyses, statistical significance was set at two-tailed p<0.05, voxel-wise corrected for familywise-error rate ($p_{FWE}<0.05$) within ROI. As default *t*-tests in SPM/SnPM provide one-tailed *p*-values, all the obtained *p*-values were multiplied by 2; it follows that, for instance, voxel-wise statistics for "FLE>CTR" or "FLE<CTR" each had to meet a more stringent statistical threshold of $p_{FWE}<0.025$ to be reported as statistically significant. For completeness, we report group differences for areas outside the above-described ROIs, if these survived a threshold of two-tailed $p_{FWE}<0.05$, corrected voxel-wise across the whole brain.

*Supplementary Figure 1: Language, WM and task-negative ROIs*

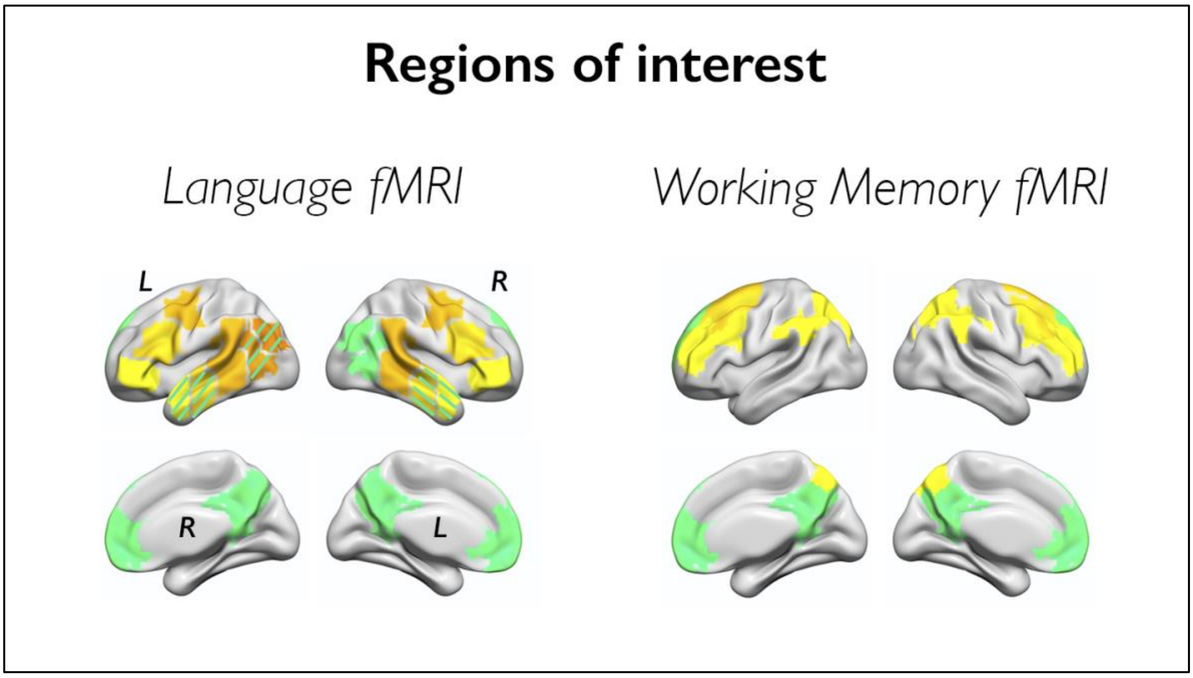


Fronto-temporo-parietal language ROIs and WM ROIs are highlighted with warm color shades. Some temporo-parietal ROIs may show task-related activation or deactivation during verbal fluency or verb generation fMRI. Such areas are depicted with warm colors and superimposed cold-colored dashed lines. Right posterior-temporal and angular language ROIs belonged to the deactivation map in both tasks; while bilateral posterior temporal/angular ROIs are used for the purpose of *p*-value correction for multiple comparisons, intergroup differences in task effects within such regions are interpreted in terms of activation or deactivation, based on each specific ROI's task-related behavior in controls. Midline task-negative ROIs, corresponding to medial DMN areas, are displayed with cold color shades.

*Voxel-based fMRI, language laterality indices*

We computed laterality indices (LIs) of frontal hemispheric dominance for language for both verbal fluency and verb generation fMRI tasks, using the word generation contrast for the former, and the "Generation minus Repetition" contrast for the latter. We used the bootstrap method and a bilateral frontal lobe mask of the SPM LI toolbox[e18] for all computations, as in prior work.[51] For demographic purposes (Table 1), we report a composite measure of frontal lobe laterality, based on a mask derived from merging the three bilateral frontal language ROIs described above (orbital inferior frontal, inferior frontal, and middle frontal gyrus). Moreover, repeat comparisons of language fMRI activation in patients with FLE and controls used frontal LIs as nuisance regressor in addition to participant age and sex (Supplementary Table 6). For these analyses, we computed three distinct frontal LI metrics, each specific to one given frontal ROI, for both verbal fluency and verb generation fMRI, using the same toolbox and method as described above. The latter LI measures are referred to as "ROI-specific" in Supplementary Table 6.

*Curves of gradient-based task-effects: group comparisons via functional data analysis*

Global differences between curves of gradient-stratified task effects were determined via functional data analysis (FDA), a branch of statistics that treats curves as functions and allows comparing these.[63] Areas between curves (AbC) were computed by summing the absolute values of group differences between β weights (*y* values) at each gradient bin (*x* values): AbC= $\Sigma_i \left| y_{group1}(x_i) - y_{group2}(x_i) \right|$. The statistical significance of differences between groups was tested using a non-parametric permutation test with 10000 permutations, as in previous work.[63] In detail, the group identity of each individual (e.g., FLE and CTR, for comparison of these two groups) was randomly reassigned without replacement, creating pseudo-groups; average curves for the two pseudo-groups were determined, and the area between these two curves, AbC', was estimated as above. Repeating this process for a number of iterations *I* (in this study, *I*=10000) led to a set of *I* AbC' values; *p*-values for the true group difference were established as the number of AbC' values greater than AbC, divided by the divided by the number of iterations *I*.

**SUPPLEMENTARY RESULTS**

*Verbal fluency fMRI: comparison of TLE and controls*

Voxel-based analyses highlighted reduced activation of left inferior frontal ($p_{FWE}$<0.05) and right mid-posterior temporal gyrus ($p_{FWE}$=0.058), and reduced deactivation of bilateral precuneus ($p_{FWE}$<0.05) in TLE compared to controls (Supplementary Figure 2). Across systems, there was lower limbic activity in TLE compared to controls at uncorrected thresholds ($p_{unc}$=0.030, *d*=-0.29). Analysis of left-hemispheric system divisions showed negative deviations across cognitive control and limbic systems ($p_{unc}$/$p_{FDR}$ =0.048/0.039; *d*=-0.26/-0.37). There were no significant differences between TLE and controls for profiles of gradient-stratified effects.

*Verb generation fMRI: comparison of TLE and controls*

In TLE, voxel-based analyses showed widespread areas of reduced activation compared to controls, which encompassed fronto-temporo-parietal and occipital cortices, with left-sided emphasis ($p_{FWE}$<0.05; Supplementary Figure 2). Across systems, there were prominent negative deviations, with the strongest effects across task positive systems, such as dorsal attention, cognitive control and salience, for analysis of systems across the whole brain (all $p_{FDR}$<0.0001; *d*=-0.60/-0.70/-0.62) and their left-hemispheric divisions (all $p_{FDR}$<0.0001; *d*=-0.63/-0.75/-0.65). There were significant differences between patients with TLE and controls for global profiles of gradient-stratified task effects (*FDA*, permuted *p*=0.046), and for effects across the majority of gradient bins (all $p_{FDR}$<0.05; *d* range= -0.60 to -0.30).

*Verbal WM fMRI: comparison of TLE and controls*

For verbal WM, there were extensive activation decreases in bilateral fronto-parietal cortices in TLE relative to controls ($p_{FWE}$<0.05; Supplementary Figure 2). Analyses across systems indicated widespread system effects, mostly involving reduced activity across dorsal attention, salience and cognitive control systems ($p_{FDR}$<0.0001/0.015/<0.001, *d*=-0.82/-0.33/-0.73). There were global differences for gradient-stratified task profiles (FDA, *p*=0.030), with reduced activity in TLE for most gradient bins ($p_{FDR}$<0.05; *d* range =-0.70 to -0.24).

*Visual WM fMRI: comparison of TLE and controls*

For the 1–0Back contrast, we found reduced deactivation of left medial prefrontal cortex in TLE compared to controls (midline anterior DMN; $p_{FWE}$<0.05; Supplementary Figure 2); there was increased activation of the superior frontal gyrus (Brodmann areas 9/46) at an exploratory threshold ($p$<0.001; peak $t$-score 3.45). Across systems, there were subthreshold effects indicating higher activity of the cognitive control system and DMN in TLE ($p_{unc}$=0.066/0.076; $d$=0.25/0.24). We found localized differences between TLE patients and controls for gradient-stratified profiles, with higher task-related signal at the transmodal apex in TLE ($p_{FDR}$<0.05, $d$=0.48 and 0.55), implying reduced deactivation.

For the 2–1Back contrast, we identified mostly right-lateralized reductions of frontoparietal activity in TLE compared to controls ($p_{FWE}$<0.05; Supplementary Figure 2). There was lower dorsal attention and cognitive control system activity in TLE, compared to healthy control data ($p_{FDR}$=0.028/0.055, $d$=-0.39/-0.33). There were no suprathreshold differences between TLE patients and controls in terms of gradient-stratified task effects.

*Supplementary Figure 2: Comparison of TLE and CTR, voxel-based analyses*

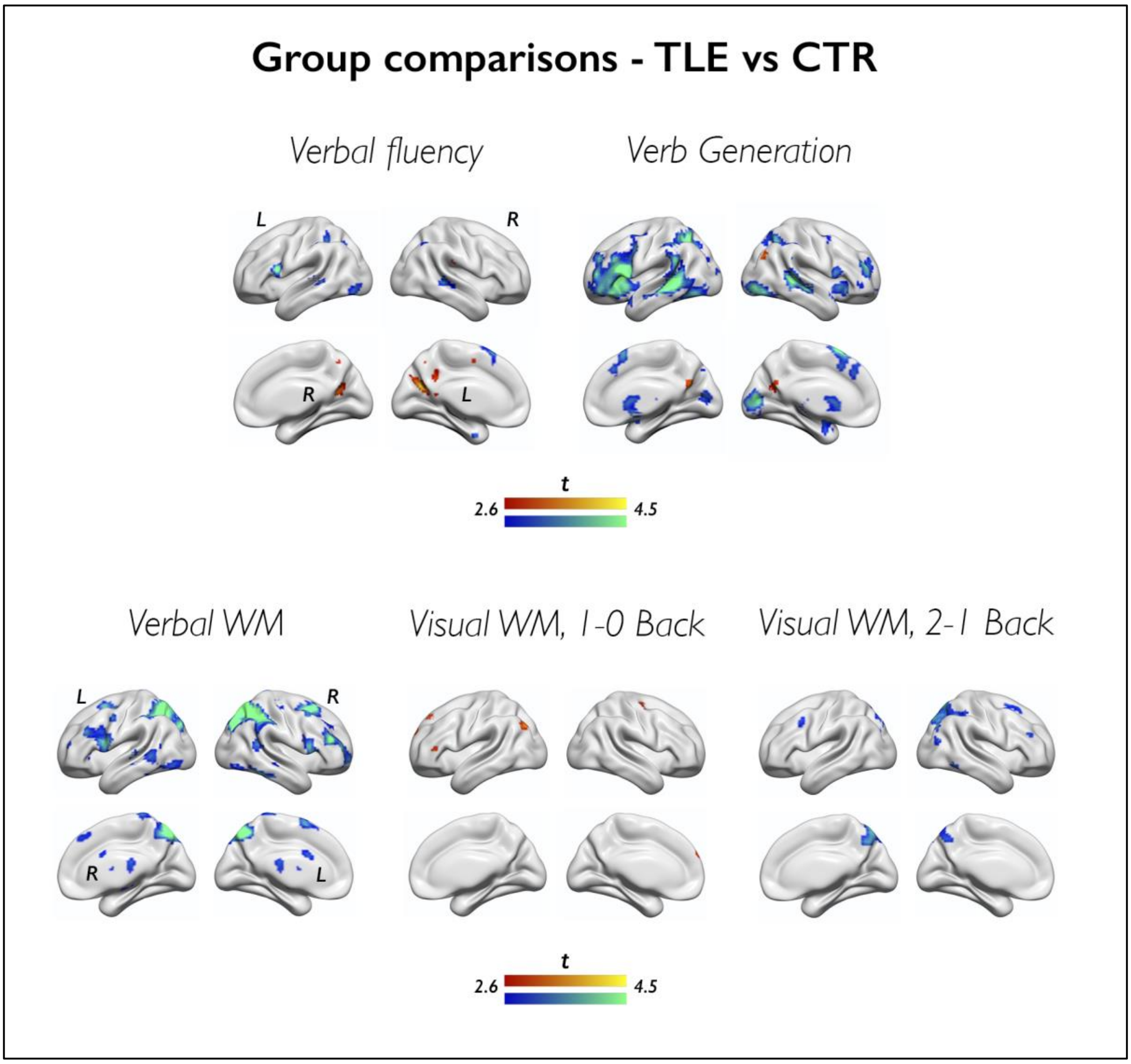


Cold/warm colors refer to lower/higher task-related effects in TLE patients than controls, respectively. For language tasks, increases in TLE exclusively map onto areas undergoing task-related deactivation (see Figure 1, main text), and are to be interpreted as areas of impaired deactivation in TLE versus controls. Group differences are shown at $p<0.005$, with an extent threshold of 10 voxels applied for display purposes; color bars indicate corresponding t-score scales. MNI coordinates and *p*-values for group comparisons are provided in the Supplementary Tables. Comparisons of TLE and healthy control participants for system and gradient analyses are shown in Figures 2 and 4 (main manuscript).

*Analyses across 17 canonical subsystems: description of system partition*

In addition to the analysis of task effects across 7 canonical functional systems, we also explored group differences using a more fine-grained 17-system parcellation, obtained from same validation dataset and via the same clustering approach as the 7-system parcellation,[36] which includes: one central and one peripheral visual system (A and B, respectively); one dorsal somatomotor system (somatomotor-A), and one system combining ventral somatomotor and auditory/posterior insular areas (somatomotor-B); two interdigitated dorsal attention and two salience subsystems (dorsal attention and salience A and B, respectively); one medial temporal and one orbital (para)limbic system divisions (limbic-A and B); three cognitive control system subsections, two of which almost exclusively encompassing lateral hemispheric areas (control-A and control-B), and one constrained to medial parietal and posterior cingulate regions (control-C); three DMN subsections, two of which encompassing both frontal and temporo-parietal regions (DMN-A, DMN-B), and one constrained to temporo-parietal regions only; and one lateral temporo-parietal system (Supplementary Figure 3). This more fine-grained system parcellation provides more refined insights in relation to task effects across executive control and default-mode systems, which are functionally heterogeneous[35,e19] and cover large patches of cortex in the 7-system partition.

*Analyses across 17 canonical subsystems: verbal fluency fMRI (Supplementary Figure 3)*

For verbal fluency in controls, we identified significant task-related effects across both salience systems ($\beta$=0.05/0.15, $p_{unc}$=0.049/$p_{FDR}$<0.001, salience-A and salience-B). Effects across cognitive control, DMN and visual systems were not homogeneous: there was significant activation across control-A, control-B, DMN-B and visual-A (central) subsystems ($\beta$=0.21/0.07/0.11/0.17, $p_{FDR}$<0.001/0.046/0.002/<0.001, respectively), and significant deactivation of control-C (medial parietal), DMN-A, DMN-C, and visual-B (peripheral) systems ($\beta$=–0.35/–0.20/–0.16/–0.10, $p_{FDR}$<0.001/<0.001/<0.001/0.046, respectively). Deactivation additionally involved the dorsal somatomotor system ($\beta$= –0.06, $p_{FDR}$=0.049).

In FLE compared to controls, we found reduced activation of cognitive control-A ($p_{FDR}$=0.017, $d$=-0.43), and reduced deactivation of cognitive control-C, DMN-A and DMN-C systems ($p_{FDR}$<0.001/0.004/0.025, $d$=0.65/0.51/0.39). In TLE compared to controls, we found reduced deactivation of cognitive control-C and DMN-C ($p_{FDR}$=0.021/0.015, $d$=0.39/0.45), and reduced activation of cognitive control-A, control-B, DMN-B and limbic-A (medial temporal) systems at uncorrected thresholds ($p_{unc}$=0.017/0.043/0.031/0.023, $d$=-0.32/-0.27/-0.29/-0.30), and. There were no differences between FLE and TLE. Overall, these findings point to an altered balance between task-related system activation and deactivation both in FLE and TLE, with slightly more prominent effects in the former.

*Analyses across 17 canonical subsystems: verb generation fMRI (Supplementary Figure 3)*

For verb generation in controls, we identified significant task-related activation across salience-A, salience-B, temporo-parietal, and peripheral visual systems ($\beta$=0.09/0.10/0.06/0.15, $p_{FDR}$=0.001/0.26/0.043/<0.001), and across control-A, control-B, and DMN-B systems ($\beta$=0.15/0.09/0.18, $p_{FDR}$<0.001/0.001/<0.001, respectively).

In FLE compared to controls, we found reduced activation of DMN-B and temporo-parietal systems at uncorrected thresholds ($p_{unc}$=0.046/0.049, *d*=-0.28/-0.27). In TLE compared to controls, we found reduced activation of virtually all those cognitive systems exhibiting positive task effects in controls, including cognitive control-A, control-B, DMN-B, temporo-parietal, salience-A and salience-B systems (all $p_{FDR}$<0.001, *d*=-0.72/-0.58/-0.71/-0.73/-0.53/-0.70, respectively; the full extent of intergroup differences is shown in Supplementary Figure 3). At uncorrected thresholds, comparison of patient groups showed stronger task-related effects across dorsal attention-A, dorsal-attention B and central visual systems in FLE compared to TLE ($p_{unc}$=0.026/0.046/0.006, *d*=-0.42/-0.37/-0.54). On balance, these findings indicate mild activation reductions in FLE, and, conversely, extensively reduced activation across most cognitive systems in TLE. Differences between patient groups mostly map onto attentional systems.

*Analyses across 17 canonical subsystems: verbal WM fMRI (Supplementary Figure 4)*

For verbal WM in controls, we identified significant task-related activation across dorsal attention-A, cognitive control-A and B, and salience-B ($\beta$=0.25/0.30/0.28/0.19, all $p_{FDR}$ <0.001), and uncorrected effects for dorsal attention B systems ($\beta$=0.05, $p_{unc}$=0.045). Task-related deactivation encompassed DMN-C, limbic A (medial temporal), and both somatomotor systems ($\beta$=–0.10/–0.05/–0.06/–0.11, all $p_{FDR}$ 0.007/0.029/0.031/<0.001), with uncorrected effects on peripheral visual system ($\beta$=–0.09, $p_{unc}$=0.035).

In FLE compared to controls, we found reduced activation of dorsal attention-A, cognitive control-A and control-B systems ($p_{FDR}$<0.001/<0.001/0.027, *d*=-0.74/-0.62/-0.40), and of dorsal attention-B and central visual systems at uncorrected thresholds ($p_{unc}$=0.026/0.031, *d*=-0.32/-0.31). Moreover, there was reduced deactivation of DMN-C in FLE compared to controls at uncorrected thresholds ($p_{unc}$=0.016, *d*=0.34). In TLE compared to controls, we found reduced activation of dorsal attention-A and B, cognitive control-A, control-B and control-C, DMN-B, temporo-parietal, central visual and salience-B systems ($p_{FDR}$<0.001/<0.001/<0.001/<0.001/<0.001/0.029/0.008/0.023/<0.001, *d*=-0.90/-0.53/-0.84/-0.72/-0.55/-0.32/-0.40/-0.34/-0.49), and stronger deactivation of the dorsal somatomotor system ($p_{FDR}$=0.031, *d*=-0.31). At uncorrected thresholds, comparison of patient groups showed lower activity of cognitive control-C and limbic-B (orbitofrontal) systems in TLE than FLE ($p_{unc}$=0.036/0.050, *d*=-0.39/-0.36). On balance, these findings point to marked activation reduction across attentional and executive control systems in both FLE and TLE, with slightly more marked alterations in TLE.

*Analyses across 17 canonical subsystems: visual WM fMRI (Supplementary Figure 4)*

In controls, the 1–0Back contrast highlighted significant activation across dorsal attention-A, cognitive control-A, control-B, and salience-B systems (β=0.15/0.12/0.10/0.06, $p_{FDR}$ <0.001/<0.001/<0.001/0.013), and deactivation of DMN-A, DMN-B, DMN-C, visual B, somatomotor-A and B systems (β=–0.10/–0.05/–0.08/–0.09/–0.07/–0.10, $p_{FDR}$<0.001/0.043/0.011/0.032/0.002/<0.001). Contrasting high versus low WM demand (2–1Back contrast) showed increasing activation across the same systems detailed above, i.e., dorsal attention-B, cognitive control-A, control-B, salience-B, and cognitive control-C (β=0.10/0.12/0.13/0.09/0.06, $p_{FDR}$=0.003/<0.001/<0.001/0.003/0.043); there was also increasing deactivation of somatomotor-A and B systems (β=–0.05/–0.07, $p_{FDR}$=0.043/0.001). In FLE compared to controls, we found increased activation of dorsal attention-A, cognitive control-A, B and C for the 1-0Back contrast ($p_{FDR}$=0.0495/0.0495/0.005/0.0495, *d*=0.35/0.38/0.55/0.38), and reduced deactivation of all three DMN subsystems (DMN-A, -B,-C; $p_{FDR}$=0.0495/0.044/0.0495, *d*=0.35/0.44/0.37), and of limbic-B (orbitofrontal) and central visual systems at uncorrected thresholds ($p_{unc}$=0.035/0.046, *d*=0.32/0.30). In TLE compared to controls, we found increased activation of cognitive control-B ($p_{unc}$=0.029, *d*=0.30) and reduced deactivation of DMN-A systems ($p_{unc}$=0.041, *d*=0.27), both at uncorrected thresholds, for the 1-0Back contrast. For the 2-1Back contrast, we found significantly reduced activation across all attentional and cognitive control systems in FLE compared to controls ($p_{FDR}$<0.001/<0.001/<0.001/0.002/<0.001/0.038/0.021, *d*=-0.66/-0.53/-0.68/-0.54/-0.56/-0.34/-0.40, for dorsal attention A and B, control-A, B and C, and salience-A and B; the full extent of intergroup differences is shown in Supplementary Figure 3). For the same task contrast, we also found reduced activation of dorsal attention-A, cognitive control-A, B and C in TLE compared to controls ($p_{FDR}$=0.002/0.033/0.040/0.033, *d*=-0.58/-0.39/-0.36/-0.38). For both 1-0Back and 2-1Back task contrasts, there were no differences between TLE and FLE. On balance, these results point to (i) enhanced activation of attentional and executive control systems and reduced deactivation of default-mode subsystems for lower WM task demands; this is followed by (ii) defective additional recruitment of attentional and executive control systems for higher task difficulty levels in both patient groups, with more marked deviations noted for FLE.

*Supplementary Figure 3: analyses across 17 systems, language fMRI*

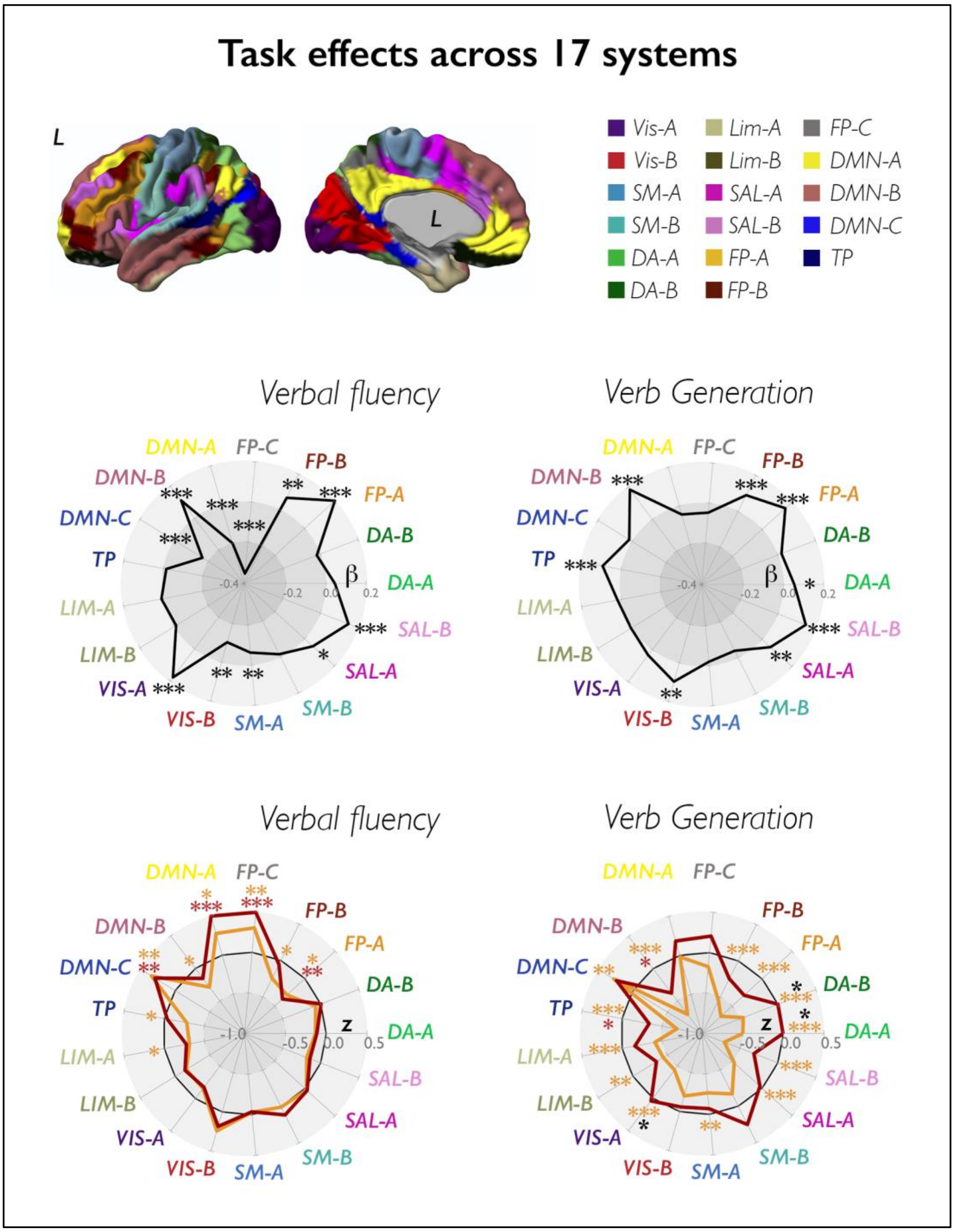


The 17 functional system parcellation, as per (Yeo et al., 2011), is displayed in the upper panel. The spider plots in the middle panels show mean task-related effects in controls, parameterized as β weights (contrast estimates) across 17 functional systems. The spider plots in the lower panels show Z-score analyses of task-related effects across the same systems; across panels, black heptagons display effects in controls (Z-score equal to zero, at each system), while effects in FLE and TLE are shown in dark red and orange lines and in correspondingly colored asterisks, respectively; ***, $p_{FDR}<0.01$; **, $p_{FDR}<0.05$; *, uncorrected $p<0.05$. Beta value for control-A task effects in controls, verbal fluency= 0.21; for display purposes, the spider plot axes reach a maximum of β= 0.20. DA: dorsal attention; FP: cognitive (frontoparietal) control;

DMN: default-mode network; TP: temporo-parietal; LIM: limbic; VIS: visual; SM: somatomotor; SAL: salience (ventral attention).

*Supplementary Figure 4: analyses across 17 systems, WM fMRI*

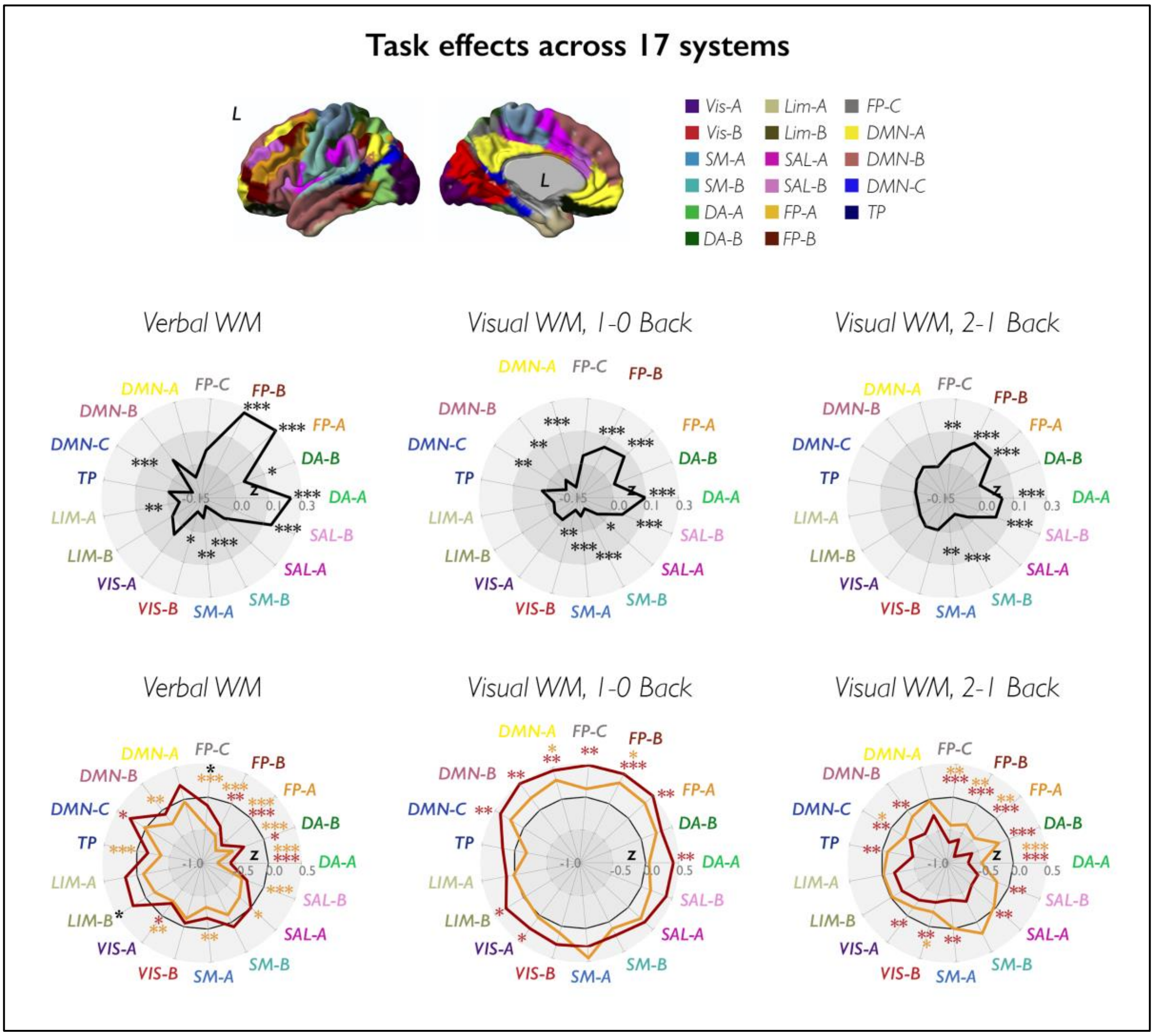


The 17 functional system parcellation, as per (Yeo et al., 2011), is displayed in the upper panel. The spider plots in the middle panels show mean task-related effects in controls, parameterized as β weights (contrast estimates) across 17 functional systems. The spider plots in the lower panels show Z-score analyses of task-related effects across the same systems; across panels, black heptagons display effects in controls (Z-score equal to zero, at each system), while effects in FLE and TLE are shown in dark red and orange lines and in correspondingly colored asterisks, respectively; ***, $p_{FDR}<0.01$; **, $p_{FDR}<0.05$; *, uncorrected $p<0.05$. Z-score for control-A/control-B/DMN-B task effects in FLE, 1-0Back visual working memory= 0.52/0.66/0.53, respectively; for display purposes, the spider plot axes reach a maximum of Z=0.5. DA: dorsal attention; FP: cognitive (frontoparietal) control; DMN: default-mode network; TP: temporo-parietal; LIM: limbic; VIS: visual; SM: somatomotor; SAL: salience (ventral attention).

*Sensitivity analyses: FLE-FCD versus controls, language fMRI*

Final comparisons included 11/12 patients with FLE-FCD and 49/47 controls for verbal fluency/verb generation, respectively (see above for quality control and participant exclusion criteria). For verbal fluency fMRI, patients with FLE and focal cortical dysplasia (FLE-FCD) had lower activation of the orbital inferior frontal gyrus (peak $t$=3.69, $p_{FWE}$<0.05, ROI-based; Supplementary Figure 5). Reduced deactivation of anterior DMN areas was found at an exploratory threshold ($t$=3.39, uncorrected $p$=0.0014). For verb generation fMRI, FLE-FCD patients had reduced activation of inferior frontal gyrus ($t$=4.69, $p_{FWE}$<0.05 within ROI), along with reduced deactivation of the right angular gyrus ($t$=4.22, $p_{FWE}$<0.05 within ROI; Supplementary Figure 5); lesser deactivation of posterior DMN areas (precuneus) was found at an exploratory threshold (uncorrected peak $t$=3.32, $p$=0.0026). Across systems, there were no significant deviations of task-related effects in FLE-FCD from control data, for both verbal fluency and verb generation fMRI. For verbal fluency, gradient analyses identified global disorganization of task-related recruitment (*FDA*, $p$=0.007), with lower task signal across intermediate bins at uncorrected thresholds ($p_{unc}$ range=0.023 to 0.036, $d$ range=-0.82 to -0.71) and qualitative trends showing increases at the transmodal apex ($p_{unc}$>0.05, $d$ range=0.23-0.33). For verb generation, there was a tendency for a positive deviation from controls at the transmodal apex ($p_{unc}$=0.046, $d$=0.67).

Overall, comparison of the FLE-FCD sample to controls broadly recapitulated the findings reported for the analysis of the whole FLE subgroup: reduced activation in frontal regions, particularly during verb generation, and reduced deactivation mapping onto midline DMN or angular regions at the voxel level. As in comparisons of the main FLE group against controls, deviations in gradient-based profiles of task effects were more evident for verbal fluency.

*Sensitivity analyses: FLE-FCD versus controls, WM fMRI*

Final comparisons included 10/11 patients with FLE-FCD and 50/49 controls for verbal and visual WM contrasts, respectively (see above for quality control and participant exclusion criteria). For verbal WM fMRI, we identified marked, bilateral frontoparietal activation reductions in FLE-FCD compared to controls ($t$-score range: 3.62-5.08, all $p_{FWE}$<0.05 within ROI; Supplementary Figure 5). As for visual WM, the 1-0Back contrast highlighted increased right dorsolateral frontal and parietal activation, and reduced deactivation of medial frontal areas in DMN territories in FLE-FCD patients compared to controls ($t$-score range: 4.20-4.37, all $p_{FWE}$<0.05 within ROI; Supplementary Figure 5). Contrasting higher and lower WM loads via the 2-1Back contrast revealed failure to elicit additional activation of bilateral dorsolateral frontal and right parietal regions ($t$-score range: 3.90-4.68; all $p_{FWE}$<0.05, ROI-based; Supplementary Figure 5). For analysis of systems, there was reduced activation of dorsal attention and cognitive control systems in FLE-FCD compared to controls for verbal WM ($p_{FDR}$=0.007/0.014, $d$=-1.52/-1.01). For the 1-0Back visual WM contrast, mean task effects were higher across cognitive control system and DMN, though $p$-values did not reach statistical significance ($p_{unc}$=0.063/0.17, $d$=0.65/0.50); for the 2-1Back visual WM contrast, there was lower task activation of the dorsal attention network ($p_{unc}$=0.049, $d$=-0.67). For verbal WM, there were global differences in gradient-stratified curves of task effects between FLE-FCD and controls (*FDA*, $p$=0.013); analyses across gradient bins showed negative deviations from controls that encompassed intermediate to transmodal gradient segments ($p_{FDR}$ range: 0.003 to 0.048; $d$ range=-1.40 to -0.8). For visual WM, there were global differences in gradient-stratified curves of task effects between FLE-FCD and controls for the 2-1Back contrast (*FDA*, $p$=0.041), and trends for the 1-0Back contrast (*FDA*, $p$=0.097). Analyses across gradient bins did not reach statistical significance for either contrast; we did observe a pattern of increased task-effects across the intermediate and transmodal gradient sections for the 1-0Back contrast ($d$ range=0.44 to 0.56), and decreased task effects across the same sections, for the 2-1Back contrast ($d$ range=-0.64 to -0.41).

On balance, comparison of the FLE-FCD sample to controls broadly recapitulated the findings obtained in analyses comparing the whole FLE subgroup to controls: (i) enhanced frontoparietal activation and reduced DMN deactivation occurring for low-level WM demands, (ii) markedly reduced activation of frontoparietal WM areas for higher task loads (2-Back), and (iii) global disorganization of task-related recruitment, evidenced via gradient-based analyses, particularly for higher task difficulty levels.

*Supplementary Figure 5: Comparison of FLE-FCD and CTR, voxel-based analyses*

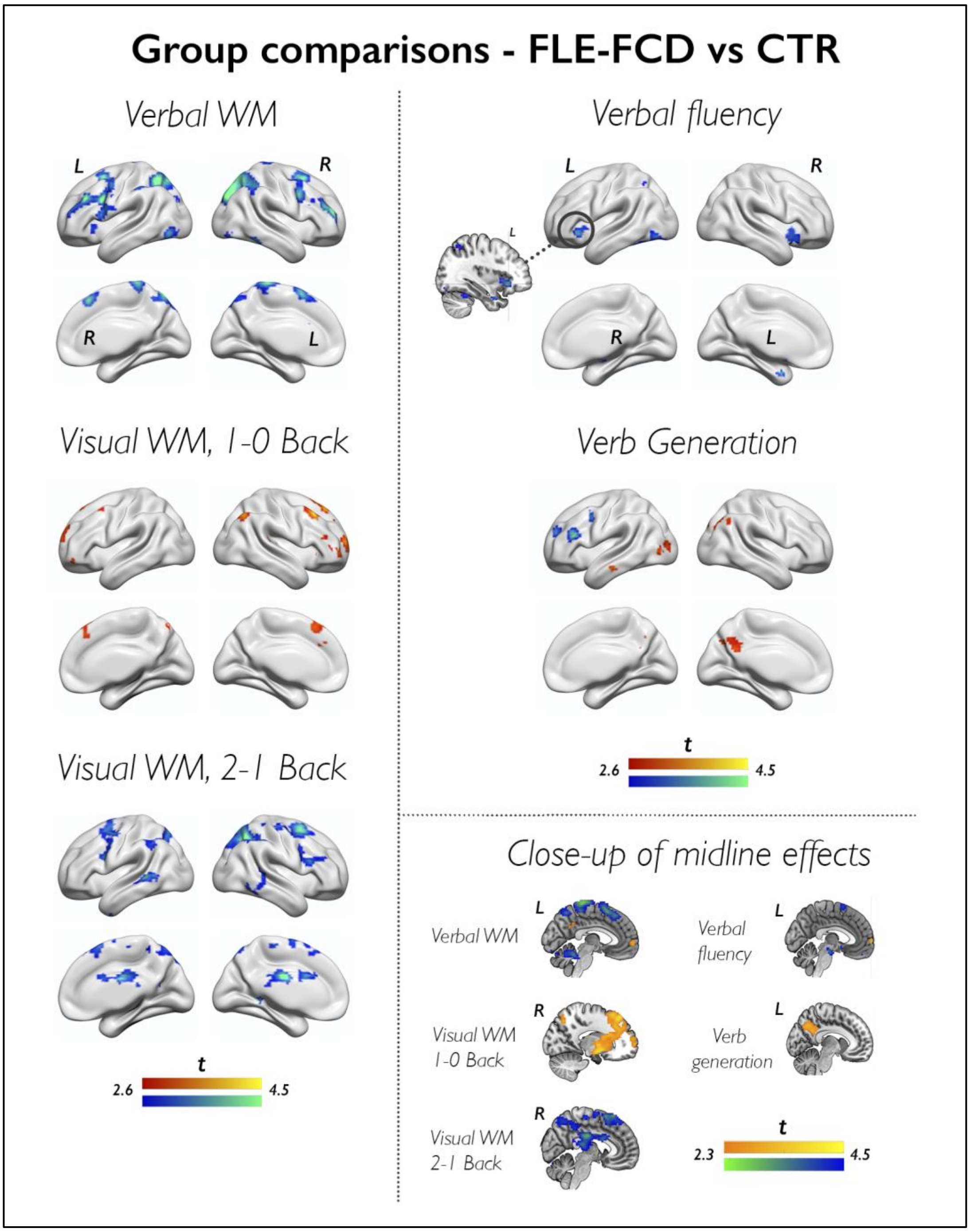


The figure shows voxel-based maps of group differences between patients with FLE-FCD and healthy controls across all the analyzed task contrasts. For brain renders, voxel-based differences are shown for the whole brain at $p<0.005$ with an extent threshold of 10 voxels applied for display purposes. Cold/warm colors refer to lower/higher task-related effects in FLE-FCD patients than controls, respectively; color bars indicate t-score scales. MNI coordinates and p-values for group comparisons within prespecified regions of interest are provided in text above. Sagittal brain slices in the lower right-hand panel ("Close up of midline effects") further illustrate group differences for effects in midline and para-midline brain areas at a more liberal statistical threshold. WM; working memory.

*Supplementary Figure 6: Analysis of left and right FLE subgroups*

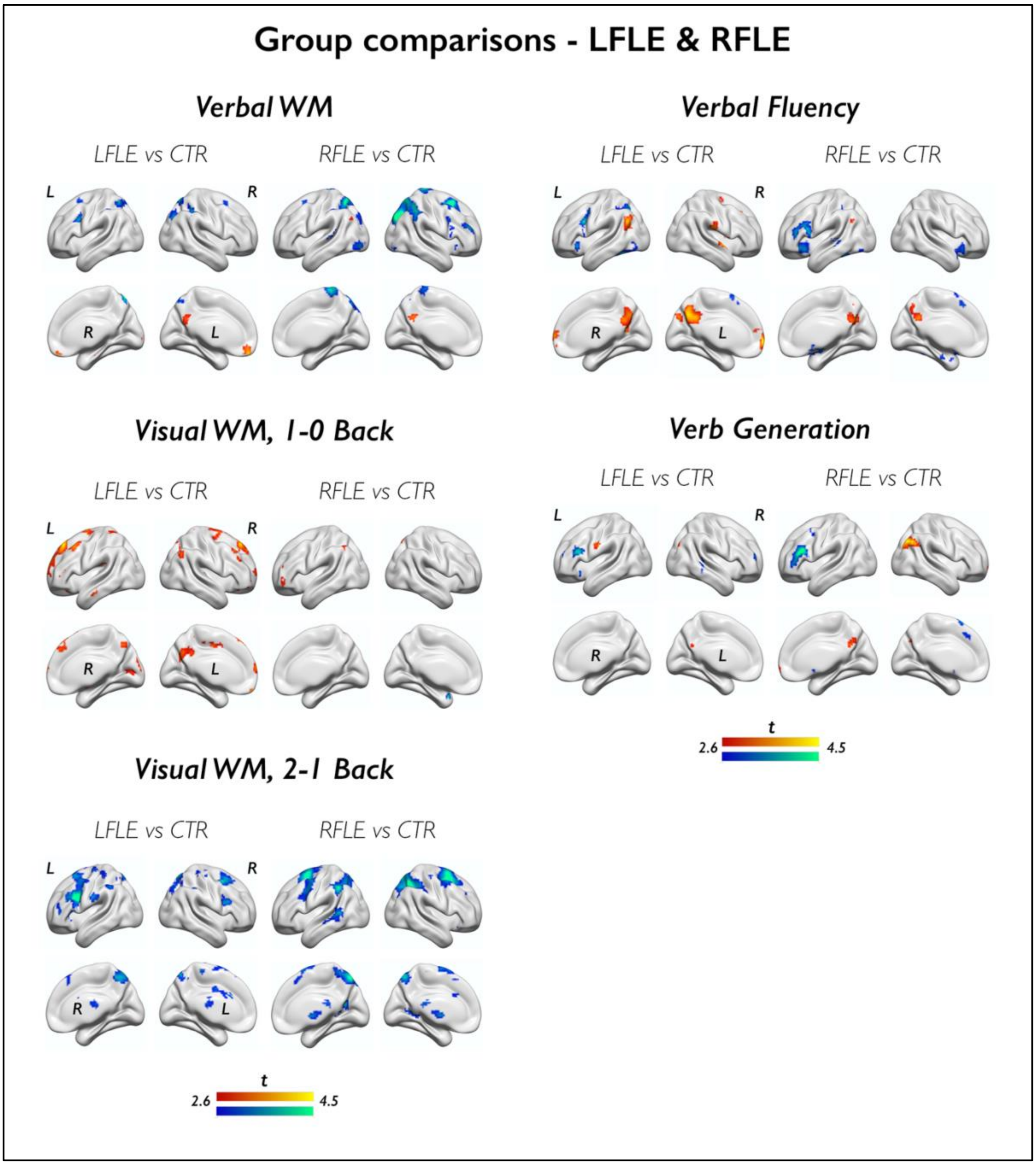


The figure shows voxel-based maps of group differences between patients with FLE-FCD and healthy controls across all the analyzed task contrasts. For brain renders, voxel-based differences are shown for the whole brain at $p<0.005$ with an extent threshold of 10 voxels applied for display purposes. Cold/warm colors refer to lower/higher task-related effects in left FLE or right FLE compared to controls controls, respectively; color bars indicate t-score scales. WM: working memory.

*Correlations of fMRI measures and cognitive performance: language fMRI*

Here, we provide a detailed report of analyses probing associations between language fMRI measures and cognitive performance across all participants. A succinct description is provided in the main manuscript text. Statistical details are provided in Supplementary Table 7.

For verbal fluency fMRI, higher letter fluency scores were associated with stronger activation of the inferior frontal gyrus at exploratory thresholds (peak $t$-score=3.56, $p$<0.001), and with stronger deactivation of the left precuneus ($p_{FWE}$<0.05, ROI-based). Similarly, higher naming scores were positively associated with stronger inferior frontal and lateral temporal activation at exploratory thresholds ($p$<0.001; peak $t$-scores=3.36 and 3.79), with lower activation of the right inferior frontal gyrus ($p$<0.001, peak $t$-score= 3.63), and with stronger deactivation of left precuneus ($p_{FWE}$<0.05, ROI-based) and left angular gyrus ($p$<0.001; peak $t$-score=3.39). There were no suprathreshold associations for analyses of systems; follow-up tests identified: (i) negative correlations between activity of the cognitive control-C subsystem, encompassing posterior cingulate cortex and precuneus, and both letter fluency and naming scores ($r_{perm}$=-0.22/-0.20, $p$=0.007/0.019), and (ii) a negative correlation between activity across the peripheral visual system and letter fluency scores only ($r_{perm}$=-0.17, $p$=0.029). Negative correlations between letter fluency scores and gradient-stratified task effects during verbal fluency were evident for bins located at the transmodal apex ($r_{perm}$=-0.17/-0.19, $p$=0.036/0.026 for significant bins); there were no significant correlations between naming scores and gradient-stratified task effects.

For verb generation fMRI, higher category fluency scores were associated with stronger activation of the left inferior frontal gyrus ($p_{FWE}$<0.05, ROI-based), and of the left middle-posterior temporal gyrus at exploratory thresholds ($p$<0.001; peak $t$-score=3.68). Similarly, higher naming scores were positively associated with stronger inferior frontal, orbital inferior frontal, middle-posterior and posterior temporal activation (all $p_{FWE}$<0.05, ROI-based), with stronger left anterior temporal activation at exploratory thresholds ($p$<0.001; peak $t$-score=3.46), and with stronger deactivation of the right angular gyrus ($p_{FWE}$<0.05, ROI-based). Across canonical systems, there was a significant positive correlation between limbic activity and naming scores ($r_{perm}$=0.16, $p$=0.046); there were no significant correlations between category fluency and task effects during verb generation. Follow-up analyses detected significant correlations between activity of the DMN-B system, encompassing lateral fronto-temporal cortices, and both category fluency and naming scores ($r_{perm}$=0.16/0.17, $p$=0.048/0.036, respectively); naming scores also correlated with task activity across the medial-temporal (limbic A) subsystem ($r_{perm}$=0.16, $p$=0.049). For gradient-based profiles, there was a near-significant association between naming scores and task effects at the transmodal apex ($r_{perm}$=-0.16, $p$=0.055); there were no significant associations between category fluency scores and verb generation task effects on gradients.

*Correlations of fMRI measures and cognitive performance: WM fMRI*

Here, we provide a detailed report of analyses probing associations between language fMRI measures and cognitive performance across all participants. A succinct description is provided in the main manuscript text. Statistical details are provided in Supplementary Table 8.

For verbal WM, higher digit span scores significantly correlated with stronger activation of bilateral frontal (all $p_{FWE}$<0.05, ROI-based), bilateral parietal areas (all $p_{FWE}$<0.05, ROI-based), and with effects in right medial prefrontal and right medial parietal cortex (all $p_{FWE}$<0.05, ROI-based). Higher 2-Back verbal WM scores were associated with stronger activation of bilateral frontal (all $p_{FWE}$<0.05, ROI-based) and bilateral parietal areas (all $p_{FWE}$<0.05, ROI-based), and with stronger deactivation of the right precuneus at an exploratory threshold ($p$<0.001/$p_{FWE}$=0.054 peak $t$-score= 3.48) Across canonical systems, activity across dorsal attention and cognitive control systems was significantly associated with digit span scores ($r_{perm}$=0.31/0.33, both $p_{FDR}$=0.0007) and verbal 2-Back scores ($\rho_{perm}$=0.23/0.21, $p_{FDR}$=0.020/0.027). As for gradient analyses, activity across intermediate and transmodal gradient sections was positively associated with both digit span ($p_{FDR}$<0.05, $r_{perm}$ range: 0.24 to 0.31) and verbal 2-Back scores ($p_{FDR}$<0.05, $\rho_{perm}$ range: 0.20 to 0.23).

For visual WM, higher visual 2-Back scores significantly correlated with stronger activation, as assessed via the 2-1Back contrast, of bilateral frontal areas (all $p_{FWE}$<0.05, ROI-based), bilateral parietal areas (all $p_{FWE}$<0.05, ROI-based), as well as with areas located in right medial prefrontal and right medial parietal cortex (all $p_{FWE}$<0.05, ROI-based). Activity across dorsal attention and cognitive control systems for the 2-1Back contrast was significantly associated with visual 2-Back scores ($\rho_{perm}$=0.37/0.39, $p_{FDR}$=<0.0001/0.0001). As for gradient analyses focusing on the 2-1Back contrast, task-related effects in the intermediate gradient section were significantly associated with visual 2-Back scores ($p_{FDR}$<0.05, $\rho_{perm}$ range: 0.20 to 0.36).

*Correlations of fMRI measures with age and onset and disease duration: FLE*

Here, we provide MNI coordinates and statistical details for the voxel-wise permutation-based regression analyses testing the associations between functional imaging patterns and disease characteristics in FLE, which are summarized in the main manuscript text. Significant associations for analyses of systems and gradients and related statistics are provided in the main manuscript text. All statistics refer to 2-tailed significance.

For verbal fluency fMRI, longer disease duration was associated with higher task effects in a rostral left middle frontal area (x=-39, y=35, z=46, $t$=5.04, $p_{FWE}$=0.0416, corrected voxel-wise across the whole brain), which bordered the anterior margin of the task-related activation map in FLE; higher effects in this area may potentially reflect compensatory recruitment. The association between longer disease duration and lesser deactivation of posterior DMN areas (left precuneus) was near-significant (x=-3, y=73, z=46, $t$=3.74, $p_{FWE}$=0.056, ROI-based). For verb generation fMRI, longer disease duration was associated with lesser deactivation of the left angular and right posterior temporal gyrus (x=-51, y=-70, z=37, $t$=3.72, $p_{FWE}$=0.0462 and x=57, y=-58, z=43, $t$=4.29, $p_{FWE}$=0.0098, respectively; both ROI-based).

For verbal WM, longer disease duration was associated with lower left and right parietal activation (x=-36, y=-49, z=37, $t$=4.55, $p_{FWE}$=0.0138 and x=-42, y=-55, z=31, $t$=4.29, $p_{FWE}$=0.0274; two independent left-peaks, ROI-based; x=36, y=-43, z=37, $t$=4.22, $p_{FWE}$=0.0314); at an exploratory thresholds, earlier age at epilepsy onset was also associated with lower bilateral parietal activation (x=-33, y=-76, z=46, $t$=4.10, $p$=0.0004, left peak; and x=48, y=-64, z=43, $t$=3.87, $p$=0.0002, right peak). At an exploratory threshold, earlier age at onset and longer disease duration were both associated with lower dorsolateral frontal activation (x=-48, y=8, z=28, $t$=3.91, $p$=0.0002, left peak, age at onset; x=-45, y=14, y=49 and x=-39, y=14, z=46, $t$=3.72/3.67, $p$=0.0004/0.0002, left peaks, duration).

For visual WM fMRI, longer epilepsy duration was associated with lower right dorsolateral frontal activation for the 1-0Back contrast at an exploratory threshold (x=30, y=17, z=49, $t$=3.72, $p$=0.0003).

*Correlations of fMRI measures with age and onset and disease duration: TLE*

Here, we report associations between task effects and disease characteristics in TLE across all three scales. All statistics refer to 2-tailed significance.

For verbal fluency fMRI, earlier age at epilepsy onset was associated with lower left orbital inferior frontal activation (x=-33, y=29, z=-17, $t$=3.82, $p_{FWE}$=0.0488, ROI-based). There were no significant associations for verb generation fMRI. For WM fMRI, longer epilepsy duration was associated with lesser deactivation of bilateral posterior cingulate cortex/precuneus for the 1-0 Back visual WM contrast (posterior DMN ROI; x=-6, y=-43, z=7, and x=-3, y=-46, z=4, $t$=3.93/3.74, $p_{FWE}$=0.0224/0.0362, left peaks; x=6, y=-46, z=7, $t$=4.03, $p_{FWE}$=0.0168, right peak). There were no significant associations for verbal WM fMRI. For analysis of systems, there was a trend-level association of lesser limbic system deactivation with age at epilepsy onset ($r_{perm}$=0.23, $p$=0.077). There were no significant associations between disease characteristics and task effects on the principal gradient.

### SUPPLEMENTARY TABLES

*<u>Supplementary Table 1. Verbal fluency fMRI</u>*

*Group comparisons: MNI coordinates and test statistics*

| ***Region*** | **MNI coordinates (x y z)** | | | ***T* score** | ***P* value (FWE)** | **MNI coordinates (x y z)** | | | ***T* score** | ***P* value (FWE)** |
|---|---|---|---|---|---|---|---|---|---|---|
| | ***Left hemisphere*** | | | | | ***Right hemisphere*** | | | | |
| **FLE < CTR** | | | | | | | | | | |
| *Inferior frontal gyrus, orbitalis* | -33 | 26 | -8 | 4.22 | 0.0104 | | | | | |
| *Inferior frontal gyrus* | -39 | 11 | 25 | 4.53 | 0.0048 | | | | | |
| | -51 | 14 | 13 | 3.9 | 0.0344 | | | | | |
| *Middle frontal gyrus* | -51 | 5 | 46 | 4.08 | 0.0146 | | | | | |
| *Middle-anterior temporal* | -63 | -28 | -5 | 4.1 | 0.0104 | | | | | |
| *Middle-posterior temporal* | -63 | -28 | 1 | 3.93 | 0.0354 | | | | | |
| *Inferior temporal/fusiform gyrus^* | -45 | -52 | -20 | 4.74 | 0.0220^ | | | | | |
| *Putamen^* | -18 | 8 | -2 | 4.77 | 0.0204^ | | | | | |
| *Cerebellum^* | | | | | | 30 | -61 | -32 | 5.45 | 0.0018^ |
| **FLE > CTR** | | | | | | | | | | |
| *Anterior temporal* | | | | | | 57 | -1 | -2 | 3.97 | 0.0174 |
| *Posterior temporal* | -51 | -61 | 28 | 3.99 | 0.0172 | | | | | |
| *Angular* | -54 | -64 | 31 | 3.92 | 0.0214 | | | | | |
| | -54 | -70 | 25 | 3.75 | 0.0338 | | | | | |
| | -45 | -76 | 40 | 3.69 | 0.0392 | | | | | |
| *Anterior task-negative ROI* | -3 | 65 | 1 | 4.12 | 0.0134 | 6 | 68 | 16 | 4.00 | 0.0194 |
| *Posterior task-negative ROI* | -9 | -55 | 34 | 4.59 | 0.0034 | 0 | -55 | 31 | 4.63 | 0.0032 |
| | -3 | -67 | 37 | 4.58 | 0.0034 | | | | | |
| **TLE < CTR** | | | | | | | | | | |
| *Inferior frontal gyrus* | -54 | 14 | 16 | 4.40 | 0.0076 | | | | | |
| **TLE > CTR** | | | | | | | | | | |
| *Posterior DMN* | -21 | -67 | 22 | 4.51 | 0.0028 | 18 | -61 | 19 | 3.55 | 0.0492 |
| **FLE < TLE** | | | | | | | | | | |
| *Cerebellum^* | | | | | | 30 | -58 | -32 | 4.94 | 0.0116^ |
| **FLE > TLE** | | | | | | | | | | |
| *Middle-posterior temporal (within de- activation map)* | -54 | -55 | 40 | 3.86 | 0.0418 | | | | | |
| *Anterior task-negative ROI* | | | | | | 18 | 47 | 46 | 3.59 | 0.0436 |

Abbreviations: CTR= controls; DMN= default mode network; FLE= patients with frontal lobe epilepsy; FWE= family-wise error; MNI= Montreal Neurological Institute; ROI= region of interest; TLE= patients with temporal lobe epilepsy. Coordinates of fMRI activation differences are provided in MNI space. The table report statistics associated with permutation-based two-tailed $t$-tests, conducted with age and sex as covariates of no interest, and 10000 permutations; $p$-values are voxel-wise FWE-corrected for multiple comparisons within prespecified regions of interest (see Supplementary Figure 1 and accompanying text), unless otherwise stated. All the reported *p*-values refer to 2-tailed statistical significance. ^area outside of prespecified ROIs, with voxel-based statistic surviving two-tailed $p_{FWE}$ <0.05, corrected across the whole brain.

### *Supplementary Table 2. Verb generation fMRI*

#### *Group comparisons: MNI coordinates and test statistics*

| ***Region*** | **MNI coordinates (x y z)** | | | ***T* score** | ***P* value (FWE)** | **MNI coordinates (x y z)** | | | ***T* score** | ***P* value (FWE)** |
|---|---|---|---|---|---|---|---|---|---|---|
| | ***Left hemisphere*** | | | | | ***Right hemisphere*** | | | | |
| **FLE < CTR** | | | | | | | | | | |
| *Inferior frontal gyrus* | -45 | 26 | 19 | 5.41 | 0.0002 | | | | | |
| *Middle frontal gyrus* | -45 | 5 | 43 | 3.55 | 0.0486 | | | | | |
| **FLE > CTR** | | | | | | | | | | |
| *Angular* | | | | | | 36 | -85 | 34 | 4.59 | 0.0024 |
| **TLE < CTR** | | | | | | | | | | |
| *Inferior frontal gyrus* | -51 | 17 | 13 | 5.55 | 0.0004 | | | | | |
| *Inferior frontal gyrus, orbitalis* | -36 | 23 | 7 | 4.68 | 0.0006 | | | | | |
| | -45 | 17 | -8 | 4.64 | 0.0008 | | | | | |
| | -33 | 32 | -2 | 4.60 | 0.0010 | | | | | |
| *Anterior temporal* | | | | | | 48 | 11 | -14 | 3.88 | 0.0206 |
| *Middle-anterior temporal* | -66 | -25 | -8 | 4.57 | 0.0028 | 51 | -25 | -11 | 4.57 | 0.0028 |
| | -54 | -10 | -17 | 3.77 | 0.0304 | | | | | |
| *Middle-posterior temporal* | -54 | -40 | -5 | 6.17 | 0.0002 | 54 | -40 | 4 | 4.65 | 0.0024 |
| | -60 | -49 | 13 | 4.45 | 0.0066 | 51 | -28 | -8 | 4.50 | 0.0056 |
| | -60 | -46 | 25 | 4.00 | 0.0232 | | | | | |
| | -51 | -40 | 40 | 4.15 | 0.0160 | | | | | |
| *Posterior temporal* | -63 | -58 | 13 | 4.19 | 0.0122 | 48 | -79 | -5 | 3.68 | 0.0456 |
| | -51 | -76 | -5 | 3.70 | 0.0438 | | | | | |
| | -54 | -70 | -8 | 3.66 | 0.0484 | | | | | |
| *Angular* | -27 | -64 | 37 | 4.08 | 0.0126 | | | | | |
| *Superior parietal lobule^* | -33 | -64 | 58 | 4.99 | 0.0112^ | | | | | |
| *Hippocampus^* | -27 | 23 | -11 | 4.82 | 0.0192^ | | | | | |
| | -33 | 32 | 2 | 4.60 | 0.0424^ | | | | | |
| *Globus pallidus^* | | | | | | 9 | 2 | -8 | 4.74 | 0.0250^ |
| **TLE > CTR** | | | | | | | | | | |
| *Angular* | -42 | -79 | 40 | 3.96 | 0.0184 | 45 | -76 | 37 | 4.59 | 0.0022 |
| *Posterior task-negative* | 12 | -61 | 22 | 3.65 | 0.0458 | | | | | |
| **FLE < TLE** | | | | | | | | | | |
| *No significant differences* | | | | | | | | | | |
| **FLE > TLE** | | | | | | | | | | |
| *Posterior temporal* | -48 | -79 | -5 | 3.84 | 0.0240 | | | | | |
| *Angular* | -21 | -79 | 34 | 3.85 | 0.0238 | 30 | -76 | 28 | 4.00 | 0.0136 |
| | -27 | -94 | 19 | 3.73 | 0.0346 | | | | | |
| *Posterior task negative ROI* | -18 | -79 | 37 | 3.88 | 0.0148 | 21 | -76 | 37 | 3.65 | 0.0338 |
| *Occipital pole^* | | | | | | 0 | -100 | 4 | 4.99 | 0.0092^ |
| *Lateral occipital cortex^* | | | | | | 39 | -91 | -2 | 4.73 | 0.0212^ |

Abbreviations: CTR= controls; DMN= default mode network; FLE= patients with frontal lobe epilepsy; FWE= family-wise error; MNI= Montreal Neurological Institute; ROI= region of interest; TLE= patients with temporal lobe epilepsy. Coordinates of fMRI activation differences are provided in MNI space. The table report statistics associated with permutation-based two-tailed *t*-tests, conducted with age and sex as covariates of no interest, and 10000 permutations; *p*-values are voxel-wise FWE-corrected for multiple comparisons within prespecified regions of interest (see Supplementary Figure 1 and accompanying text), unless otherwise stated. All the reported *p*-values refer to 2-tailed statistical significance. ^area outside of prespecified ROIs, with voxel-based statistic surviving two-tailed $p_{FWE}$ <0.05, corrected across the whole brain.

### *Supplementary Table 3. Verbal WM fMRI*

#### *Group comparisons: MNI coordinates and test statistics*

| ***Region*** | **MNI coordinates (x y z)** | | | ***T* score** | ***P* value (FWE)** | **MNI coordinates (x y z)** | | | ***T*-score** | ***P* value (FWE)** |
|---|---|---|---|---|---|---|---|---|---|---|
| | ***Left hemisphere*** | | | | | ***Right hemisphere*** | | | | |
| **FLE < CTR** | | | | | | | | | | |
| *FEF/premotor ROI (BA 6-8)* | -27 | 8 | 58 | 3.80 | 0.0392 | 27 | 11 | 55 | 5.00 | 0.0008 |
| *Lateral prefrontal ROI (BA 9/46)* | | | | | | 36 | 11 | 49 | 4.00 | 0.0176 |
| *Dorsal Parietal ROI* | -33 | -58 | 55 | 5.26 | 0.0006 | 18 | -67 | 61 | 5.56 | 0.0004 |
| | -15 | -70 | 55 | 4.07 | 0.0248 | 30 | -79 | 37 | 5.34 | 0.0004 |
| | -45 | -49 | 58 | 4.03 | 0.0274 | 42 | -46 | 46 | 4.98 | 0.0012 |
| | | | | | | 51 | -46 | 34 | 4.38 | 0.0088 |
| *Posterior task-negative ROI* (area within activation map)* | | | | | | 24 | -76 | 34 | 4.55 | 0.0014 |
| **FLE > CTR** | | | | | | | | | | |
| *Anterior task-negative ROI* | 0 | 47 | -20 | 4.25 | 0.0046 | | | | | |
| *Posterior task-negative ROI* | -9 | -55 | 31 | 3.96 | 0.014 | | | | | |
| **TLE < CTR** | | | | | | | | | | |
| *FEF/premotor ROI (BA 6-8)* | -33 | 2 | 67 | 4.53 | 0.0052 | 27 | 17 | 58 | 4.89 | 0.0016 |
| | -27 | 11 | 58 | 3.95 | 0.0276 | 27 | 5 | 70 | 4.15 | 0.0154 |
| *Lateral prefrontal ROI (BA 9/46)* | -54 | 11 | 19 | 4.15 | 0.0128 | 45 | 41 | 22 | 4.69 | 0.0018 |
| | | | | | | 36 | 11 | 49 | 3.93 | 0.0230 |
| | | | | | | 33 | 53 | 10 | 3.91 | 0.0244 |
| | | | | | | 27 | 23 | 52 | 3.85 | 0.0312 |
| | | | | | | 30 | 41 | 43 | 3.74 | 0.0430 |
| *Dorsal Parietal ROI* | -27 | -61 | 49 | 6.10 | 0.0002 | 21 | -67 | 61 | 6.28 | 0.0002 |
| | -6 | -67 | 52 | 5.36 | 0.0004 | 42 | -46 | 40 | 5.98 | 0.0002 |
| | -45 | -55 | 58 | 4.82 | 0.0014 | 30 | -79 | 37 | 5.91 | 0.0002 |
| | -24 | -82 | 37 | 4.15 | 0.0186 | | | | | |
| *Posterior task-negative ROI* (area within activation map)* | -6 | -76 | 46 | 3.63 | 0.0288 | 9 | -76 | 46 | 4.28 | 0.0032 |
| | -12 | -67 | 43 | 3.50 | 0.0446 | 24 | -76 | 34 | 3.94 | 0.0114 |
| | | | | | | 3 | -55 | 49 | 3.60 | 0.0318 |
| *Inferior temporal gyrus^* | | | | | | 57 | -49 | -20 | 4.75 | 0.0136^ |
| *Superior occipital gyrus^* | -27 | -82 | 34 | 4.44 | 0.0434^ | | | | | |
| **TLE > CTR** | | | | | | | | | | |
| *No significant differences* | | | | | | | | | | |
| **FLE < TLE** | | | | | | | | | | |
| *No significant differences* | | | | | | | | | | |
| **FLE > TLE** | | | | | | | | | | |
| *Posterior task-negative ROI (area within deactivation map)* | -3 | -49 | 13 | 3.51 | 0.0486 | 6 | -46 | 13 | 4.27 | 0.0062 |

Abbreviations: BA= Brodmann Area; CTR= controls; DMN= default mode network; FEF= frontal eye field; FLE= patients with frontal lobe epilepsy; FWE= family-wise error; MNI= Montreal Neurological Institute; ROI= region of interest; TLE= patients with temporal lobe epilepsy. Coordinates of fMRI activation differences are provided in MNI space. The table report statistics associated with permutation-based two-tailed *t*-tests, conducted with age and sex as covariates of no interest, and 10000 permutations; *p*-values are voxel-wise FWE-corrected for multiple comparisons within prespecified regions of interest (see Supplementary Figure 1 and accompanying text), unless otherwise stated. All the reported *p*-values refer to 2-tailed statistical significance. ^area outside of prespecified ROIs, with voxel-based statistic surviving two-tailed $p_{FWE}$ <0.05, corrected across the whole brain. * a small portion of the dorsal precuneus, which formally belongs to the posterior task-negative ROI, displays task-related activation during WM.

*Supplementary Table 4. Visual WM fMRI, 1-0 Back*

*Group comparisons: MNI coordinates and test statistics*

| ***Region*** | **MNI coordinates (x y z)** | | | ***T* score** | ***P* value (FWE)** | **MNI coordinates (x y z)** | | | ***T* score** | ***P* value (FWE)** |
|---|---|---|---|---|---|---|---|---|---|---|
| | ***Left hemisphere*** | | | | | ***Right hemisphere*** | | | | |
| **FLE < CTR** | | | | | | | | | | |
| *No significant differences* | | | | | | | | | | |
| **FLE > CTR** | | | | | | | | | | |
| *FEF/premotor ROI (BA 6-8)* | -45 | 23 | 43 | 3.76 | 0.0444 | | | | | |
| *Lateral prefrontal ROI (BA 9/46)* | -48 | 20 | 43 | 3.85 | 0.0388 | | | | | |
| *Dorsal Parietal ROI* | | | | | | 24 | -76 | 55 | 4.11 | 0.0242 |
| *Anterior task-negative ROI* | -18 | 62 | 13 | 3.62 | 0.0454 | | | | | |
| **TLE < CTR** | | | | | | | | | | |
| *No significant differences* | | | | | | | | | | |
| **TLE > CTR** | | | | | | | | | | |
| *Anterior task-negative ROI* | -12 | 59 | 28 | 3.63 | 0.0364 | | | | | |
| | -6 | 68 | 13 | 3.60 | 0.0400 | | | | | |
| **FLE < TLE** | | | | | | | | | | |
| *No significant differences* | | | | | | | | | | |
| **FLE > TLE** | | | | | | | | | | |
| *No significant differences* | | | | | | | | | | |

Abbreviations: BA= Brodmann Area; CTR= controls; DMN= default mode network; FEF= frontal eye field; FLE= patients with frontal lobe epilepsy; FWE= family-wise error; MNI= Montreal Neurological Institute; ROI= region of interest; TLE= patients with temporal lobe epilepsy. Coordinates of fMRI activation differences are provided in MNI space. The table report statistics associated with permutation-based two-tailed *t*-tests, conducted with age and sex as covariates of no interest, and 10000 permutations; the associated *p*-values are voxel-wise FWE-corrected for multiple comparisons within pre-defined regions of interest (see Supplementary Figure 1 and accompanying text). All the reported *p*-values refer to 2-tailed statistical significance.

### <u>*Supplementary Table 5. Visual WM fMRI, 2-1 Back*</u>

### *Group comparisons: MNI coordinates and test statistics*

| ***Region*** | **MNI coordinates (x y z)** | | | ***T* score** | ***P* value (FWE)** | **MNI coordinates (x y z)** | | | ***T* score** | ***P* value (FWE)** |
|---|---|---|---|---|---|---|---|---|---|---|
| | ***Left hemisphere*** | | | | | ***Right hemisphere*** | | | | |
| **FLE < CTR** | | | | | | | | | | |
| *FEF/premotor ROI (BA 6-8)* | -24 | 8 | 58 | 4.37 | 0.0052 | 30 | 11 | 58 | 5.23 | 0.0010 |
| | -39 | 5 | 61 | 4.28 | 0.0068 | 6 | 26 | 64 | 3.62 | 0.0454 |
| | -12 | -1 | 67 | 4.02 | 0.0136 | | | | | |
| *Lateral prefrontal ROI (BA 9/46)* | -39 | 14 | 25 | 4.64 | 0.0018 | 27 | 23 | 52 | 3.99 | 0.0132 |
| | -42 | 2 | 46 | 3.74 | 0.0290 | 39 | 8 | 52 | 3.83 | 0.0232 |
| | | | | | | 36 | 5 | 31 | 3.63 | 0.0384 |
| *Dorsal Parietal ROI* | -15 | -70 | 58 | 4.65 | 0.0034 | 12 | -67 | 55 | 4.79 | 0.0022 |
| | -51 | -46 | 40 | 4.40 | 0.0066 | 18 | -67 | 64 | 4.73 | 0.0024 |
| | -36 | -49 | 37 | 3.95 | 0.0260 | 51 | -43 | 49 | 4.00 | 0.0218 |
| | -24 | -61 | 43 | 3.91 | 0.0288 | 36 | -49 | 37 | 3.77 | 0.0450 |
| *Posterior task-negative ROI* (area within activation map)* | | | | | | 3 | -61 | 46 | 3.61 | 0.0464 |
| **FLE > CTR** | | | | | | | | | | |
| *No significant differences* | | | | | | | | | | |
| **TLE < CTR** | | | | | | | | | | |
| *FEF/premotor ROI (BA 6-8)* | | | | | | 30 | 11 | 58 | 3.99 | 0.0142 |
| *Dorsal Parietal ROI* | | | | | | 30 | -82 | 43 | 4.37 | 0.0076 |
| | | | | | | 12 | -76 | 58 | 4.28 | 0.0096 |
| | | | | | | 18 | -64 | 58 | 3.97 | 0.0232 |
| | | | | | | 27 | -70 | 49 | 3.74 | 0.0494 |
| *Posterior task-negative ROI* (area within activation map)* | | | | | | 3 | -64 | 43 | 3.67 | 0.0338 |
| | | | | | | 9 | -79 | 46 | 3.58 | 0.0424 |
| **TLE > CTR** | | | | | | | | | | |
| *No significant differences* | | | | | | | | | | |
| **FLE < TLE** | | | | | | | | | | |
| *No significant differences* | | | | | | | | | | |
| **FLE > TLE** | | | | | | | | | | |
| *No significant differences* | | | | | | | | | | |

Abbreviations: BA= Brodmann Area; CTR= controls; DMN= default mode network; FEF= frontal eye field; FLE= patients with frontal lobe epilepsy; FWE= family-wise error; MNI= Montreal Neurological Institute; ROI= region of interest; TLE= patients with temporal lobe epilepsy. Coordinates of fMRI activation differences are provided in MNI space. The table report statistics associated with permutation-based two-tailed *t*-tests, conducted with age and sex as covariates of no interest, and 10000 permutations; the associated *p*-values are voxel-wise FWE-corrected for multiple comparisons within pre-defined regions of interest (see Supplementary Figure 1 and accompanying text). All the reported *p*-values refer to 2-tailed statistical significance. * a small portion of the dorsal precuneus, which formally belongs to the posterior task-negative ROI, displays task-related activation during WM.

*Supplementary Table 6. Language fMRI: frontal group differences covaried for language LI*

*Group comparisons, frontal lobe ROIs: MNI coordinates and test statistics*

| ***FRONTAL ROI*** | **MNI coordinates (x y z)** | | | ***T* score** | ***P* value (FWE)** | **MNI coordinates (x y z)** | | | ***T* score** | ***P* value (FWE)** |
|---|---|---|---|---|---|---|---|---|---|---|
| | ***Left hemisphere*** | | | | | ***Right hemisphere*** | | | | |
| **FLE < CTR, *Verbal fluency fMRI*** | | | | | | | | | | |
| *Inferior frontal gyrus (covaried for ROI-specific LI)* | -39 | 11 | 25 | 3.86 | 0.0404 | | | | | |
| *Inferior frontal gyrus, orbitalis (covaried for ROI-specific LI)* | -30 | 26 | -8 | 4.29 | 0.0078 | 36 | 17 | -5 | 3.93 | 0.0240 |
| *Middle frontal gyrus (covaried for ROI-specific LI)* | -48 | 5 | 43 | 3.82 | 0.0348 | | | | | |
| **FLE < CTR, *Verb Generation fMRI*** | | | | | | | | | | |
| *Inferior frontal gyrus (covaried for ROI-specific LI)* | -45 | 23 | 19 | 4.46 | 0.0050 | | | | | |

Abbreviations: CTR= controls; FLE= patients with frontal lobe epilepsy; FWE= family-wise error; LI= laterality index; MNI= Montreal Neurological Institute; ROI= region of interest. Coordinates of fMRI activation differences are provided in MNI space. These repeat group comparisons focused on differences in frontal lobe areas only (see main text). We used LI measures specific to each of the three frontal lobe ROIs; i.e., comparisons of orbital inferior frontal activation used task-specific LI measures computed using the orbital inferior frontal ROI, and so forth. All group comparisons were conducted via two-tailed *t*-tests, using ROI-specific LI, age and sex as covariates of no interest; the associated *p*-values are voxel-wise FWE-corrected for multiple comparisons within each region of interest (see Supplementary Figure 1 and accompanying text). All the reported *p*-values refer to 2-tailed statistical significance.

### Supplementary Table 7. Language fMRI and cognitive test scores

*Group comparisons: MNI coordinates and test statistics for multiple regressions*

| ***Region*** | **MNI coordinates (x y z)** | | | ***T* score** | ***P* value (FWE)** | **MNI coordinates (x y z)** | | | ***T* score** | ***P* value (FWE)** |
|---|---|---|---|---|---|---|---|---|---|---|
| | ***Left hemisphere*** | | | | | ***Right hemisphere*** | | | | |
| ***VF fMRI & letter fluency - positive*** | | | | | | | | | | |
| *No corrected associations** | | | | | | | | | | |
| ***VF fMRI & letter fluency - negative*** | | | | | | | | | | |
| *Posterior task-negative ROI* | -6 | -61 | 52 | 3.99 | 0.0184 | 12 | -61 | 52 | 3.64 | 0.0488 |
| ***VF fMRI & naming - positive*** | | | | | | | | | | |
| *No corrected associations** | | | | | | | | | | |
| ***VF fMRI & naming - negative*** | | | | | | | | | | |
| *Posterior task-negative ROI* | -9 | -79 | 40 | 4.03 | 0.0140 | | | | | |
| | -9 | -61 | 49 | 3.63 | 0.0408 | | | | | |
| ***VG fMRI & category fluency - positive*** | | | | | | | | | | |
| *Inferior frontal gyrus* | -48 | 29 | 22 | 4.19 | 0.0108 | | | | | |
| *Inferior frontal gyrus, orbitalis* | -30 | 32 | -2 | 3.95 | 0.0174 | | | | | |
| ***VG fMRI & category fluency - negative*** | | | | | | | | | | |
| *No corrected associations** | | | | | | | | | | |
| ***VG fMRI & naming - positive*** | | | | | | | | | | |
| *Inferior frontal gyrus* | -51 | 20 | 16 | 4.44 | 0.0028 | | | | | |
| | -42 | 8 | 25 | 4.03 | 0.0134 | | | | | |
| *Inferior frontal gyrus, orbitalis* | -27 | 29 | -5 | 3.81 | 0.0112 | | | | | |
| | -42 | 29 | -11 | 3.46 | 0.0318 | | | | | |
| *Middle-posterior temporal* | -48 | -43 | -11 | 3.95 | 0.0230 | | | | | |
| | -54 | -40 | 1 | 3.67 | 0.0484 | | | | | |
| *Posterior temporal* | -45 | -52 | 19 | 3.78 | 0.0278 | | | | | |
| | -45 | -58 | -2 | 3.57 | 0.0498 | | | | | |
| ***VG fMRI & naming - negative*** | | | | | | | | | | |
| *Angular* | | | | | | 42 | -79 | 34 | 4.36 | 0.0040 |
| | | | | | | 33 | -88 | 34 | 3.96 | 0.0124 |
| | | | | | | 54 | -67 | 34 | 3.66 | 0.0342 |

Abbreviations: CTR= controls; DMN= default mode network; FLE= patients with frontal lobe epilepsy; FWE= family-wise error; MNI= Montreal Neurological Institute; ROI= region of interest; TLE= patients with temporal lobe epilepsy; VF=verbal fluency; VG= verb generation. Permutation-based multiple regression analyses probing associations between fMRI activity and cognitive test scores were conducted using age, sex and group allocation as covariates of no interest, and 10000 permutations; the associated *p*-values are voxel-wise FWE-corrected for multiple comparisons within predefined regions of interest (see Supplementary Figure 1). All the reported *p*-values refer to 2-tailed statistical significance. *Associations between fronto-temporal activation during verbal fluency fMRI and letter fluency or naming scores were evident at uncorrected thresholds only (x=-51, y=26, z=28, $t$=3.56, $p_{FWE}$=0.108, $p_{unc}$=0.0004, left IFG, verbal fluency fMRI and letter fluency; x=-48, y=8, z=22, $t$=3.36, $p_{FWE}$=0.135, $p_{unc}$=0.0008, left IFG, verbal fluency fMRI and naming; x=-54, y-37, z=1, $t$=3.79, $p_{FWE}$=0.056, $p_{unc}$=0.0001, middle-posterior temporal, verbal fluency fMRI and naming).

*Supplementary Table 8. WM fMRI and cognitive test scores*

*Group comparisons: MNI coordinates and test statistics for multiple regressions*

| ***Region*** | **MNI coordinates (x y z)** | | | ***T* score** | ***P* value (FWE)** | **MNI coordinates (x y z)** | | | ***T*-score** | ***P* value (FWE)** |
|---|---|---|---|---|---|---|---|---|---|---|
| | ***Left hemisphere*** | | | | | ***Right hemisphere*** | | | | |
| ***Verbal WM fMRI &digit span - positive*** | | | | | | | | | | |
| *FEF/premotor ROI (BA 6-8)* | -33 | 5 | 64 | 5.25 | 0.0002 | 27 | 14 | 64 | 5.92 | 0.0002 |
| | -24 | 11 | 55 | 4.58 | 0.0040 | 42 | 38 | 34 | 4.83 | 0.0014 |
| | -42 | 26 | 34 | 4.30 | 0.0104 | 48 | 29 | 37 | 4.51 | 0.0046 |
| | -3 | 26 | 55 | 3.820 | 0.0426 | 24 | 32 | 40 | 3.84 | 0.0396 |
| *Lateral prefrontal ROI (BA 9/46)* | -42 | 11 | 34 | 4.70 | 0.0026 | 36 | 11 | 46 | 5.18 | 0.0002 |
| | -42 | 29 | 28 | 4.43 | 0.0062 | 39 | 5 | 34 | 4.64 | 0.0032 |
| | -51 | 23 | 31 | 4.28 | 0.0108 | 39 | 41 | 34 | 4.97 | 0.0004 |
| | -33 | 56 | 13 | 4.22 | 0.0124 | 27 | 23 | 52 | 4.66 | 0.0026 |
| | -45 | 5 | 52 | 3.97 | 0.0256 | 27 | 32 | 40 | 4.10 | 0.0168 |
| | | | | | | 33 | 56 | 1 | 4.96 | 0.0004 |
| *Dorsal Parietal ROI* | -30 | -64 | 52 | 5.47 | 0.0004 | 33 | -67 | 49 | 6.35 | 0.0002 |
| | -48 | -49 | 58 | 3.95 | 0.0326 | 27 | -82 | 46 | 5.80 | 0.0002 |
| *Anterior task-negative ROI (within activation map)** | 15 | 38 | 16 | 3.66 | 0.0350 | | | | | |
| *Posterior task-negative ROI (within activation map)** | -6 | -76 | 46 | 3.55 | 0.0482 | 9 | -73 | 43 | 4.31 | 0.0050 |
| | | | | | | 24 | -76 | 34 | 3.77 | 0.0242 |
| | | | | | | 3 | -55 | 49 | 3.64 | 0.0384 |
| *Dorsal parietal/dorsal precuneus (outside prespecified ROI, within activation map)^* | -6 | -70 | 52 | 4.88 | 0.0134^ | 15 | -67 | 64 | 5.16 | 0.0042^ |
| | -15 | -82 | 49 | 4.70 | 0.0254^ | 9 | -64 | 52 | 4.81 | 0.0178^ |
| *Cerebellum^* | -6 | -46 | -20 | 5.22 | 0.0034^ | 9 | -40 | -35 | 5.03 | 0.0064^ |
| | | | | | | 6 | -46 | -20 | 5.01 | 0.0076^ |
| *Frontopolar cortex^* | | | | | | 21 | 62 | -8 | 4.58 | 0.0346^ |
| ***Verbal WM fMRI &digit span - negative*** | | | | | | | | | | |
| *No suprathreshold associations* | | | | | | | | | | |
| ***Verbal WM fMRI & verbal 2Back - positive*** | | | | | | | | | | |
| *FEF/premotor ROI (BA 6-8)* | -33 | 26 | 31 | 3.73 | 0.0406 | | | | | |
| *Lateral prefrontal ROI (BA 9/46)* | -39 | 5 | 37 | 4.72 | 0.0010 | 36 | 41 | 13 | 4.18 | 0.0066 |
| | -33 | 5 | 31 | 4.59 | 0.0014 | 36 | 11 | 40 | 3.62 | 0.0392 |
| | -33 | 17 | 25 | 4.35 | 0.0030 | | | | | |
| *Dorsal Parietal ROI* | -64 | 52 | -64 | 3.83 | 0.0434 | 36 | -49 | 37 | 4.19 | 0.0142 |
| | | | | | | 36 | -58 | 40 | 4.15 | 0.0156 |
| | | | | | | 36 | -43 | 37 | 4.06 | 0.0212 |
| | | | | | | 42 | -46 | 40 | 3.82 | 0.0458 |
| *(Dorsal parietal, outside ROI boundaries)^* | | | | | | 36 | -46 | 34 | 4.61 | 0.0336^ |
| *Inferior frontal gyrus^* | -33 | 14 | 22 | 4.80 | 0.0204^ | | | | | |
| ***Verbal WM fMRI & verbal 2Back - negative*** | | | | | | | | | | |
| *No suprathreshold associations* | | | | | | | | | | |
| ***2-1 Back visual WM & visual 2Back - positive*** | | | | | | | | | | |
| *FEF/premotor ROI (BA 6-8)* | -42 | 26 | 34 | 5.20 | 0.0002 | 33 | 14 | 58 | 5.90 | 0.0002 |
| | -27 | 8 | 64 | 4.89 | 0.0004 | 24 | 35 | 43 | 4.28 | 0.0048 |
| | -12 | 14 | 61 | 4.05 | 0.0098 | 0 | 26 | 52 | 4.90 | 0.0004 |

| | | | | | | | | | | |
|---|---|---|---|---|---|---|---|---|---|---|
| | -15 | -7 | 67 | 3.56 | 0.0458 | 42 | 35 | 28 | 4.56 | 0.0010 |
| | | | | | | 48 | 23 | 31 | 4.12 | 0.0080 |
| *Lateral prefrontal ROI (BA 9/46)* | -42 | 29 | 31 | 5.42 | 0.0004 | 33 | 56 | 4 | 5.40 | 0.0004 |
| | -39 | 20 | 28 | 4.98 | 0.0008 | 30 | 20 | 52 | 5.08 | 0.0008 |
| | -48 | 8 | 46 | 4.02 | 0.0138 | 42 | 41 | 25 | 4.98 | 0.0008 |
| | -39 | 50 | 16 | 3.86 | 0.0208 | 39 | 11 | 52 | 4.56 | 0.0016 |
| *Dorsal Parietal ROI* | | | | | | 3 | -64 | 49 | 7.71 | 0.0002 |
| | | | | | | 27 | -73 | 37 | 6.12 | 0.0002 |
| | | | | | | 36 | -64 | 46 | 5.89 | 0.0002 |
| *Anterior task-negative ROI (within activation map)** | -6 | 29 | 55 | 4.12 | 0.0084 | 9 | 32 | 55 | 3.94 | 0.0150 |
| | | | | | | 6 | 41 | 46 | 3.90 | 0.0170 |
| | | | | | | 12 | 35 | 46 | 3.60 | 0.0364 |
| | | | | | | 18 | 38 | 40 | 3.51 | 0.0492 |
| *Posterior task-negative ROI (within activation map)** | -12 | -67 | 43 | 4.63 | 0.0018 | 3 | -61 | 46 | 7.22 | 0.0002 |
| | -6 | -76 | 46 | 3.89 | 0.0214 | 6 | -76 | 46 | 5.37 | 0.0004 |
| | | | | | | 24 | -76 | 34 | 4.93 | 0.0006 |
| *Inferior frontal gyrus^* | -36 | 20 | -8 | 6.1 | 0.0002 | 33 | 26 | -5 | 6.72 | 0.0002 |
| *Pre-supplementary motor area^* | | | | | | 6 | 29 | 52 | 5.05 | 0.0060 |
| | | | | | | 9 | 38 | 40 | 4.39 | 0.0492 |
| *Superior occipital gyrus^* | -30 | -82 | 31 | 4.47 | 0.0380 | 6 | -7 | 7 | 4.43 | 0.0442 |
| *Thalamus^* | -15 | -4 | -8 | 4.94 | 0.0084 | | | | | |
| | -12 | -4 | 7 | 4.58 | 0.0284 | | | | | |
| *Subthalamus/midbrain^* | -3 | -16 | -23 | 5.41 | 0.0016 | | | | | |
| ***2-1 Back visual WM & visual 2 Back - negative*** | | | | | | | | | | |
| *No suprathreshold associations* | | | | | | | | | | |

Abbreviations: BA= Brodmann Area; CTR= controls; DMN= default mode network; FEF= frontal eye field; FLE= patients with frontal lobe epilepsy; FWE= family-wise error; MNI= Montreal Neurological Institute; ROI= region of interest; TLE= patients with temporal lobe epilepsy. Coordinates of voxels showing significant associations between fMRI activity and cognitive performance are provided in MNI space. All permutation-based multiple regression analyses were conducted using age, sex and group allocation as covariates of no interest; the associated *p*-values are voxel-wise FWE-corrected for multiple comparisons within predefined regions of interest (see Supplementary Figure 1 and accompanying text). All the reported *p*-values refer to 2-tailed statistical significance. ^area outside of prespecified ROIs, with voxel-based statistic surviving two-tailed $p_{FWE} <0.05$, corrected across the whole brain. * a small portion of the superior frontal gyrus and of the dorsal precuneus, which formally belong to the anterior and posterior task-negative ROIs, display task-related activation during WM.

e4. Nelson HE. National Adult Reading Test (NART): For the assessment of premorbid intelligence in patients with dementia: Test manual: Nfer-Nelson; 1982.

e5. Wechsler D. Wechsler adult intelligence scale—3rd ed.(WAIS-3®). San Antonio, TX: Harcourt Assessment; 1997.

e6. Bird CM, Papadopoulou K, Ricciardelli P, Rossor MN, Cipolotti L. Monitoring cognitive changes: psychometric properties of six cognitive tests. Br J Clin Psychol 2004; 43(Pt 2): 197-210.

e7. McKenna P, Warrington EK. Graded naming test: Manual: NFER-Nelson; 1983.

e8. Tombaugh TN. Trail Making Test A and B: normative data stratified by age and education. Arch Clin Neuropsychol 2004; 19(2): 203-14.

e9. Coughlan AK, Hollows SE. The adult memory and information processing battery (AMIPB). Psychology Department, St James Hospital, Leeds, UK 1985

e10. Caciagli L, Wandschneider B, Centeno M, Vollmar C, Vos SB, Trimmel K, *et al.* Motor hyperactivation during cognitive tasks: An endophenotype of juvenile myoclonic epilepsy. Epilepsia 2020; 61(7): 1438-52.

e11. Dale AM, Fischl B, Sereno MI. Cortical surface-based analysis. I. Segmentation and surface reconstruction. Neuroimage 1999; 9(2): 179-94.

e12. Fischl B, Sereno MI, Dale AM. Cortical surface-based analysis. II: Inflation, flattening, and a surface-based coordinate system. Neuroimage 1999; 9(2): 195-207.

e13. Greve DN, Fischl B. Accurate and robust brain image alignment using boundary-based registration. Neuroimage 2009; 48(1): 63-72.

e14. Fedorenko E, Hsieh PJ, Nieto-Castanon A, Whitfield-Gabrieli S, Kanwisher N. New method for fMRI investigations of language: defining ROIs functionally in individual subjects. J Neurophysiol 2010; 104(2): 1177-94.

e15. Chai LR, Mattar MG, Blank IA, Fedorenko E, Bassett DS. Functional Network Dynamics of the Language System. Cereb Cortex 2016; 26(11): 4148-59.

e16. Fan L, Li H, Zhuo J, Zhang Y, Wang J, Chen L, *et al.* The Human Brainnetome Atlas: A New Brain Atlas Based on Connectional Architecture. Cereb Cortex 2016; 26(8): 3508-26.

e17. Alves PN, Foulon C, Karolis V, Bzdok D, Margulies DS, Volle E, *et al.* An improved neuroanatomical model of the default-mode network reconciles previous neuroimaging and neuropathological findings. Commun Biol 2019; 2: 370.

e18. Wilke M, Lidzba K. LI-tool: a new toolbox to assess lateralization in functional MR-data. J Neurosci Methods 2007; 163(1): 128-36.

e19. Braga RM, Buckner RL. Parallel Interdigitated Distributed Networks within the Individual Estimated by Intrinsic Functional Connectivity. Neuron 2017; 95(2): 457-71.e5.
